# Heterogeneity of Treatment Effects Across Nine Glucose-Lowering Drug Classes in Type 2 Diabetes: Extension of the LEGEND-T2DM Network Study

**DOI:** 10.64898/2026.01.06.26343548

**Authors:** Hsin Yi Chen, Thomas Falconer, Anna Ostropolets, Tara V. Anand, Xinzhuo Jiang, David Dávila-García, Linying Zhang, Ruochong Fan, Hannah Morgan-Cooper, Marc A. Suchard, George Hripcsak

**Affiliations:** Department of Biomedical Informatics, Columbia University Irving Medical Center, New York, NY; Johnson C Johnson, Titusville, NJ; Institute for Informatics, Data Science and Biostatistics, Washington University in St. Louis, St. Louis, MO; Stanford School of Medicine and Stanford Health Care, Palo Alto, CA; Department of Biostatistics, Fielding School of Public Health, University of California, Los Angeles; VA Informatics and Computing Infrastructure, United States Department of Veterans Affairs

**Keywords:** Comparative effectiveness, Glucose-lowering drugs, Heterogeneity of treatment effect, Observational study, Personalized medicine, Real-world evidence

## Abstract

**Aims/Hypothesis:** Understanding heterogeneous patient responses to various glucose-lowering therapies is crucial for advancing personalized treatment approaches and optimizing outcomes for type 2 diabetes mellitus. While average treatment effects are known for many drug classes, patient responses may differ by underlying clinical and demographic factors. We hypothesize that major glucose-lowering drug classes exhibit heterogeneous treatment effects (HTE) across patient subgroups defined by key clinical and demographic characteristics.

**Methods:** This is a large-scale observational cohort study replicated in six data-sources across the Observational Health Data Sciences and Informatics network. New-user, active-comparator cohorts were constructed for patients with type 2 diabetes mellitus initiating one of the nine antihyperglycemic drug classes. Large-scale propensity score adjustment for measured confounding, empirical calibration using negative control outcomes, and random-effects meta-analysis were employed to estimate calibrated hazard ratios (HRs). HTE was assessed by comparing differences in log HRs across 10 demographic and clinical subgroups.

**Results:** Evidence of HTE was observed across hyperlipidemia, hypertension, obesity, and sex subgroups. Biguanides (vs. DPP-4i) were protective against acute myocardial infarction in patients with hyperlipidemia, and against heart failure hospitalization in patients with obesity. SGLT-2 inhibitors (vs. GLP-1 receptor agonists) reduced stroke risk only in non-obese patients. Sex-specific patterns also emerged: women taking GLP-1 receptor agonists had a higher risk of diarrhea, and women taking SGLT-2 inhibitors had a lower risk of stroke compared with DPP-4 inhibitors; these associations were not seen for male patients.

**Conclusions:** This hypothesis-generating study identified several potential signals (blood pressure status, lipid status, obesity status, and sex) where there exists treatment effect heterogeneity for several classes of type 2 diabetes mellitus drugs. These preliminary findings highlight the potential for personalized type 2 diabetes mellitus treatment recommendations based on patient characteristics.

## INTRODUCTION

Type 2 diabetes mellitus is a major cause of morbidity and mortality, affecting more than 525 million people globally [1]. Type 2 diabetes mellitus is also a major risk factor for the development of cardiovascular disease [2], chronic kidney disease [3], stroke [4], and can lead to both micro- and macrovascular complications including hepatic dysfunction, neuropathy, and retinopathy [5–7].

Therapeutic options for type 2 diabetes mellitus have expanded over the last decade with the emergence of sodium-glucose co-transporter-2 inhibitors (SGLT2i) and glucagon-like peptide-1 receptor agonists (GLP-1 RA), which reduced the risk of major cardiovascular events in randomized controlled trials (RCT) [8–11]. The recent LEGEND-T2DM study [12, 13] added evidence in our understanding of the relative effects of type 2 diabetes mellitus agents after treatment with metformin on cardiovascular risk (3 and 4-point Major Adverse Cardiac Events (MACE)) and patient-centered safety outcomes by conducting direct head-to-head comparisons of second-line antihyperglycemic agents. However, patients with type 2 diabetes mellitus are a heterogeneous group, varying widely in terms of age, sex, race, and comorbidities [14]. These factors may significantly modify the benefits and risks associated with different glucose-lowering therapies [15–17]. Already, pharmacogenetic studies have demonstrated that individual genetic variations can significantly influence drug response [18, 19], underscoring the value of investigating patient-specific factors.

While the present observational study does not directly assess genetic markers, it recognizes the importance of exploring heterogeneity of treatment effect (HTE) through readily available clinical and demographic data, which provides a complementary and highly relevant avenue for identifying differential treatment responses in real-world populations. Understanding this HTE is crucial for advancing personalized medicine and optimizing therapeutic choices to match individual patient profiles. While previous observational research has explored some aspects of HTE [20], a comprehensive assessment across a broad range of modern glucose-lowering agents and an extensive set of clinically relevant patient subgroups remains a critical unmet need.

Here, we aim to systematically investigate HTE for nine major classes of glucose-lowering drugs across a comprehensive array of predefined clinical and demographic subgroups. Specifically, we aim to determine if the comparative effectiveness and safety of these drug classes for outcomes including major adverse cardiovascular events, renal events, and other patient-centered safety endpoints, differ significantly based on patient characteristics such as demographics, diabetes severity, comorbidities, and relevant clinical history.

## METHODS

We conducted a large-scale, multinational, real-world comparative effectiveness and safety study, building upon the original LEGEND-T2DM [13] and LEGEND-HTN [21] studies. The study was approved by the Columbia University Institutional Review Board (approval date, December 2024; reference number AAAO7805).

### Data Sources

The study is an observational cohort study based on routinely collected health care data replicated across 6 databases mapped to the Observational Medical Outcomes Partnership (OMOP) Common Data Model (CDM) [22]. The analysis was performed in May 2025. The data sources varied in provenance (administrative claims, electronic health records (EHR)) and origin (the United States and Japan). Database source descriptions can be found in Supplementary Table S1.

### Drug Exposures

Pairwise class-level exposure comparisons were performed for nine commonly available classes of oral and injectable non-insulin glucose-lowering drugs: (1) Alpha-Glucosidase Inhibitors, (2) Biguanides, (3) Dipeptidyl Peptidase-4 Inhibitors (DPP-4i), (4) dual Glucose-dependent Insulinotropic Polypeptide and Glucagon-Like Peptide-1 Receptor Agonists (GLP-1 RA), (5) GLP-1 RA, (6) Meglitinides, (7) Sodium-Glucose Cotransporter-2 Inhibitors (SGLT-2i), (8) Sulfonylureas (SU), and (9) Thiazolidinediones.

For each of the nine drug classes, exposure cohorts were further characterized by concomitant metformin use at the index date (i.e., cohorts of patients initiating the study drug class with concurrent metformin, and cohorts initiating without concurrent metformin). Continuous drug exposure was defined as consecutive drug prescriptions within a 30-day window. An on-treatment (per-protocol) time-at-risk definition was used; subjects were followed from index date to treatment discontinuation.

### Study Population

The study population comprised adults (≥18 years of age) diagnosed with type 2 diabetes mellitus who initiated treatment with a drug agent from one of the nine specified glucose-lowering drug classes. The index date for each patient was the date of the first observed prescription or administration of a drug from any of these nine classes.

Study inclusion criteria include: 1) a diagnosis of type 2 diabetes mellitus on or before the index date; 2) at least 365 days of continuous observation in the database prior to the index date. Study exclusion criteria include: 1) prior diagnoses of type 1 diabetes mellitus or secondary diabetes mellitus at any time on or before the index date; 2) any exposure to a drug from the specified comparator classes (for a given pairwise comparison) prior to the index date; 3) insulin use 30 days prior to index date, unless insulin was part of a combination product with one of the index drug classes initiated on the index date.

### Clinical and Demographic Subgroups

To assess presence of heterogeneity of treatment effects, the study population within each drug exposure cohort was further stratified by several predefined clinical and demographic subgroups (Table 1). These subgroups were defined at the index date unless otherwise specified.

**Table 1.** Subgroup comparisons in this study.

| Subgroup | Comparisons |
| --- | --- |
| Age | Pairwise comparisons between:<br>• Younger: <21 years<br>• Middle-aged: 21–60 years<br>• Older: >60 years |
| Biological Sex | Comparison between Female and Male (as recorded in database based on SNOMED CT codes) |
| Renal Impairment | Stratified into three categories based on diagnosis codes for chronic kidney disease and end-stage renal disease, dialysis procedures, and relevant laboratory measures (e.g., estimated glomerular filtration rate, serum creatinine, urine albumin). We compare:<br>• No renal impairment vs Renal impairment without dialysis<br>• No renal impairment vs Renal impairment on dialysis |
| Obesity | Presence of obesity, defined as a Body Mass Index (BMI) >30 kg/m<sup>2</sup>, body weight >120 kg, or a diagnosis code for obesity. Compared against the non-obese subgroup. |
| Poorly Controlled Diabetes | Defined as an HbA1c >8% (64 mmol/mol) or a diagnosis code indicating uncontrolled type 2 diabetes or poor diabetes control. Compared against those not meeting these criteria. |
| Diabetic Ketoacidosis (DKA) | History of DKA based on diagnosis codes. Compared against those with no history of DKA. |
| Diabetic Retinopathy | History of diabetic retinopathy based on diagnosis codes. Compared against those with no history of diabetic retinopathy. |
| Essential Hypertension | History of essential hypertension based on diagnosis codes. Compared against those with no history of essential hypertension. |
| Hyperlipidemia | History of hyperlipidemia based on diagnosis codes. Compared against those with no history of hyperlipidemia. |
| Metabolic Dysfunction-Associated Steatotic Liver Disease (MASLD) | History of MASLD (including non-alcoholic fatty liver disease or non-alcoholic steatohepatitis) based on diagnosis codes. Compared against those with no history of MASLD. |

### Study Outcomes

The study assessed a range of cardiovascular effectiveness and patient-centered safety outcomes. The primary outcomes from the original LEGEND-T2DM study (composite CV endpoints of 3- and 4-point MACE) were considered alongside an expanded list of patient-centered safety outcomes, including: (1) Acute myocardial infarction, (2) Acute renal failure, (3) Hospitalization with heart failure, (4) Stroke (ischemic or hemorrhagic), (5) Abnormal weight gain, (6) Acute pancreatitis, (7) Diabetic ketoacidosis, (8) Diarrhea, (9) Hypoglycemia, (10) Vomiting, and (11) Hepatic Failure.

All outcome cohorts were constructed using previously validated phenotype algorithms, typically involving one or more diagnosis codes recorded in inpatient or outpatient settings. Drug exposure and outcome cohort definitions are from the previously conducted LEGEND-T2DM study [12, 13], while subgroup cohort definitions can be found in the Observational Health Data Sciences and Informatics (OHDSI) Phenotype Library [23].

### Statistical Analysis

The statistical approach mirrors that of the previously conducted LEGEND-T2DM and LEGEND-HTN studies [12, 13, 21]. Briefly, the statistical analysis involved a federated analysis to calculate hazards ratios across databases (conducting large scale propensity score adjustment [24] to control for both measured and indirectly measured confounding [25] from tens of thousands of baseline patient characteristics), calibrating each hazards ratio estimate using a set of 20 negative control outcomes (Supplementary Table S2) [26], then using meta-analytic methods for evidence aggregation across data sources that pass study diagnostics. The initial federated analyses were conducted using the R packages in the HADES suite [27, 28] orchestrated via Strategus [29], and the subsequent meta analyses were conducted using the R package metafor [30].

#### Study Diagnostics

While blinded to the results for each of our analyses, we performed study diagnostics to evaluate the internal and external validity of findings. These include power calculations estimating minimum detectable relative risk (MDRR), preference score (a transformation of propensity score that adjusts for prevalence differences between populations) distributions to evaluate empirical equipoise and population generalizability, patient characteristics to evaluate cohort balance before and after propensity score adjustment, and expected absolute systematic error (EASE), a summary measure of expected systematic bias. Only data sources that met predefined diagnostic thresholds [31] were included in the final analysis. Specifically, a study passed diagnostics if the MDRR was <4, >25% of subjects in each cohort had preference scores between 0.3 and 0.7, the maximum SMD <0.15 after propensity score adjustment, and EASE score <0.25.

#### Heterogeneity of Treatment Effects

Each participating site provided calibrated hazard ratio (HR) estimates for each exposure-outcome-subgroup comparison. Briefly, the hazard ratio represents the relative hazard of experiencing an outcome at time *t* between individuals receiving the target vs. comparator drug classes (Equation 1) [32].

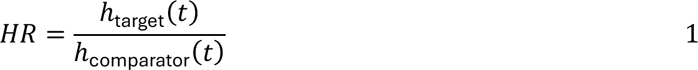

To quantify heterogeneity of treatment effects across subgroups, we calculated the difference between log-transformed hazard ratios, denoted as Δ ln(*HR*), obtained between comparator subgroups (e.g., Male vs. Female; Age <21 vs. Age 21-60; Hypertension vs. No Hypertension) within the same drug-outcome comparison (Equation 2). The corresponding standard error associated with this difference (*SE*_Δln(*HR*)_) was computed using the subgroup-specific standard errors (Equation 3). We assumed that log-transformed HR estimates follow an asymptotically normal distribution [33].

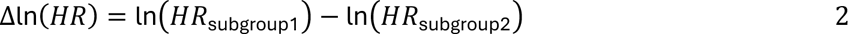

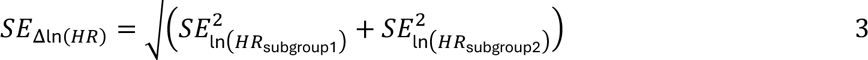

After Δln(*HR*) and *SE*_Δln(*HR*)_ were calculated, they were aggregated across the participating, non-overlapping data sources through random-effects meta-analysis [34]. This approach produced a summary effect estimate that accounted for both within-data source (sampling) variability and between-data source (heterogeneity) variability. The random-effects model assumed that the per-data source likelihoods (derived from the calibrated HRs and their standard errors) were approximately normally distributed.

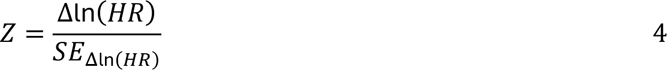

The statistical significance of subgroup differences was determined by calculating a Z-score test statistic (Equation 4) and the corresponding p-value with Type I Error rate *α* < 0.05. As this was a hypothesis-generating analysis, we ranked and reported all p-values from lowest (“most” significant) to highest and report the 5 “strongest” potential signals for HTE, regardless of whether they reached a significance threshold.

## RESULTS

Across six databases, there were a total of 3,031,892 type 2 diabetes patients that fulfilled study criteria (Table 2). Demographic descriptions, drug exposure duration, and outcome counts are reported in Tables 3-5.

**Table 2.** Cohort Sizes Across Databases. Cohort sizes among adults with type 2 diabetes mellitus (T2DM), stratified by index glucose-lowering drug class and selected demographic subgroups, across data sources. Counts are reported for the overall T2DM population, patients fulfilling study inclusion criteria, and index drug class cohorts, without excluding concomitant metformin use. Subgroup counts are shown within each index drug class cohort. For privacy protection, cells reported as “–5” indicate suppressed counts representing between 1 and 4 patients. Cohort counts including *all* index drug classes and corresponding cohorts excluding concomitant metformin use is provided in the Supplementary Materials.

| Cohort Name | CUIMC, *n* | JMDC, *n* | MDCD, *n* | MDCR, *n* | STARR OMOP, *n* | WashU, *n* |
| --- | --- | --- | --- | --- | --- | --- |
| **All T2DM Patients** | **138367** | **854819** | **873887** | **756418** | **111674** | **296727** |
| **T2DM patients fulfilling inclusion criteria** | **70189** | **782022** | **561171** | **205698** | **77421** | **129379** |
| **Biguanides** | **23087** | **41743** | **141652** | **42009** | **31321** | **4947** |
| Male | 10715 | 29092 | 53342 | 22012 | 15677 | 2930 |
| Female | 12369 | 12651 | 88310 | 19997 | 15638 | 2017 |
| Age <=20 | 242 | 394 | 11400 | 0 | 411 | 52 |
| Age 21-60 | 9861 | 34992 | 110340 | 467 | 13766 | 2414 |
| Age 61-80 | 11051 | 6357 | 19025 | 35975 | 14971 | 2164 |
| HbA1c > 8% | 685 | 0 | 0 | 0 | 541 | 13 |
| History of MASLD | 123 | 0 | 1196 | 207 | 232 | 13 |
| Renal Impairment not on Dialysis | 3247 | 6535 | 22116 | 9538 | 3500 | 551 |
| Renal Impairment on Dialysis | 121 | 52 | 619 | 130 | 162 | -5 |
| Obese | 12413 | 14059 | 81975 | 15277 | 12858 | 2457 |
| History of DKA | 308 | 732 | 2204 | 199 | 185 | 78 |
| History of Diabetic Retinopathy | 174 | 0 | 799 | 396 | 461 | 20 |
| History of Hyperlipidemia | 11379 | 24505 | 69065 | 29538 | 14321 | 2269 |
| History of Hypertension | 13626 | 19392 | 81580 | 30153 | 14591 | 2683 |
| **GLP-1 RAs** | **5896** | **1992** | **20549** | **4470** | **7858** | **1286** |
| Male | 2281 | 1264 | 5800 | 1776 | 3306 | 656 |
| Female | 3612 | 728 | 14749 | 2694 | 4550 | 630 |
| Age <=20 | 78 | 25 | 1434 | 0 | 172 | 19 |
| Age 21-60 | 3191 | 1742 | 16784 | 77 | 4155 | 775 |
| Age 61-80 | 2427 | 225 | 2281 | 4198 | 3293 | 467 |
| HbA1c > 8% | 324 | 0 | 0 | 0 | 287 | -5 |
| History of MASLD | 80 | 0 | 392 | 73 | 242 | 6 |
| Renal Impairment not on Dialysis | 1131 | 466 | 4749 | 1243 | 1593 | 176 |
| Renal Impairment on Dialysis | 67 | 26 | 161 | 13 | 194 | 10 |
| Obese | 4554 | 828 | 15424 | 2802 | 5513 | 850 |
| History of DKA | 121 | 160 | 774 | 37 | 76 | 26 |
| History of Diabetic Retinopathy | 143 | 0 | 596 | 87 | 324 | 11 |
| History of Hyperlipidemia | 3331 | 1132 | 11970 | 3054 | 4715 | 655 |
| History of Hypertension | 3519 | 932 | 12991 | 3051 | 4585 | 677 |
| **SGLT2is** | **5130** | **55685** | **15102** | **7268** | **7109** | **1241** |
| Male | 2958 | 41174 | 7201 | 4223 | 4196 | 807 |
| Female | 2172 | 14511 | 7901 | 3045 | 2911 | 434 |
| Age <=20 | -5 | 206 | 223 | 0 | 17 | -5 |
| Age 21-60 | 1792 | 44698 | 11440 | 44 | 2543 | 562 |
| Age 61-80 | 2796 | 10781 | 3280 | 5475 | 3815 | 594 |
| HbA1c > 8% | 311 | 0 | 0 | 0 | 207 | 5 |
| History of MASLD | 38 | 0 | 211 | 45 | 109 | 7 |
| Renal Impairment not on Dialysis | 1620 | 13381 | 5003 | 3058 | 2195 | 298 |
| Renal Impairment on Dialysis | 62 | 331 | 173 | 48 | 153 | 7 |
| Obese | 3171 | 20853 | 9339 | 2825 | 3272 | 665 |
| History of DKA | 75 | 639 | 409 | 47 | 61 | 10 |
| History of Diabetic Retinopathy | 146 | 0 | 423 | 109 | 245 | 9 |
| History of Hyperlipidemia | 3503 | 37607 | 9999 | 4166 | 4302 | 750 |
| History of Hypertension | 3689 | 34530 | 10724 | 4134 | 4206 | 758 |
| **Sulfonylureas** | **5792** | **4192** | **30726** | **8834** | **11276** | **1791** |
| Male | 3064 | 3072 | 12379 | 4895 | 6177 | 1089 |
| Female | 2727 | 1120 | 18347 | 3939 | 5096 | 702 |
| Age <=20 | -5 | 40 | 623 | 0 | 17 | -5 |
| Age 21-60 | 1860 | 3382 | 24771 | 97 | 4299 | 670 |
| Age 61-80 | 3049 | 770 | 4926 | 6681 | 5887 | 911 |
| HbA1c > 8% | 437 | 0 | 0 | 0 | 502 | 9 |
| History of MASLD | 19 | 0 | 292 | 31 | 93 | 7 |
| Renal Impairment not on Dialysis | 1114 | 617 | 6767 | 3330 | 2131 | 301 |
| Renal Impairment on Dialysis | 90 | 9 | 431 | 131 | 368 | 16 |
| Obese | 2621 | 1228 | 17204 | 3006 | 4310 | 760 |
| History of DKA | 65 | 55 | 794 | 119 | 94 | 30 |
| History of Diabetic Retinopathy | 92 | 0 | 721 | 302 | 357 | 7 |
| History of Hyperlipidemia | 2766 | 2323 | 17214 | 6604 | 5343 | 890 |
| History of Hypertension | 3522 | 1945 | 20029 | 7044 | 5742 | 1001 |
| **DPP4s** | **6497** | **58782** | **12514** | **5318** | **4434** | **604** |
| Male | 3144 | 42324 | 4747 | 2765 | 2187 | 317 |
| Female | 3352 | 16458 | 7767 | 2553 | 2247 | 287 |
| Age <=20 | -5 | 152 | 287 | 0 | -5 | -5 |
| Age 21-60 | 1996 | 44978 | 9373 | 53 | 1405 | 241 |
| Age 61-80 | 3484 | 13652 | 2558 | 3984 | 2431 | 289 |
| HbA1c > 8% | 610 | 0 | 0 | 0 | 235 | -5 |
| History of MASLD | 33 | 0 | 157 | 16 | 43 | -5 |
| Renal Impairment not on Dialysis | 1492 | 8797 | 3244 | 2079 | 851 | 127 |
| Renal Impairment on Dialysis | 134 | 286 | 252 | 79 | 119 | 7 |
| Obese | 2961 | 15470 | 7341 | 1759 | 1626 | 253 |
| History of DKA | 114 | 554 | 390 | 84 | 34 | 5 |
| History of Diabetic Retinopathy | 142 | 0 | 448 | 194 | 130 | 8 |
| History of Hyperlipidemia | 3522 | 36799 | 8057 | 4095 | 2228 | 318 |
| History of Hypertension | 4227 | 30964 | 8817 | 4306 | 2310 | 358 |

**Table 3.** Patient Demographics. Baseline demographic characteristics of adults with type 2 diabetes mellitus who met all study inclusion criteria. Demographic variables include age group, sex, race, and ethnicity, defined using standardized concepts within the OMOP Common Data Model. Continuous values are presented as the median and interquartile range (median [IQR]), while binary values are presented as counts and percentages (*n* [%]).

| Demographic Variable | CUIMC | JMDC | MDCD | MDCR | STARR OMOP | WashU |
| --- | --- | --- | --- | --- | --- | --- |
| **Continuous variables, median (IQR)** |  |  |  |  |  |  |
| Age in years | 64 (20) | 51 (16) | 53 (25) | 74 (12) | 63 (20) | 61 (20) |
| Observation time (days) prior to index | 3508 (5110) | 1434 (1486) | 1115 (1334) | 1077 (1139) | 2918 (3059) | 4944 (4105) |
| Observation time (days) after index | 984 (1541) | 902 (1229) | 775 (1142) | 540 (740) | 1238 (1735) | 2914664 (2067) |
| Time (days) between cohort start and end | 984 (1541) | 902 (1229) | 775 (1142) | 540 (740) | 1226 (1731) | 2914664 (2067) |
| **Binary variables, n (%)** |  |  |  |  |  |  |
| gender = MALE | 33404 (47.59%) | 461591 (59.03%) | 217809 (38.81%) | 97545 (47.42%) | 39140 (50.55%) | 62693 (48.46%) |
| gender = FEMALE | 36777 (52.4%) | 320431 (40.97%) | 343362 (61.19%) | 108153 (52.58%) | 38263 (49.42%) | 66679 (51.54%) |
| race = White | 26805 (38.19%) | NA | 282840 (50.4%) | NA | 31644 (40.87%) | 90506 (69.95%) |
| race = Black or African American | 12284 (17.5%) | NA | 176856 (31.52%) | NA | 4948 (6.39%) | 34049 (26.32%) |
| race = Asian | 2058 (2.93%) | NA | NA | NA | 17188 (22.2%) | 1763 (1.36%) |
| race = Native Hawaiian or Other Pacific Islander | NA | NA | NA | NA | 1375 (1.78%) | NA |
| ethnicity = Hispanic or Latino | 17096 (24.36%) | NA | 24329 (4.34%) | NA | 16090 (20.78%) | 1955 (1.51%) |
| ethnicity = Not Hispanic or Latino | 35723 (50.9%) | NA | NA | NA | 56333 (72.76%) | 120348 (93.02%) |

**Table 4.**
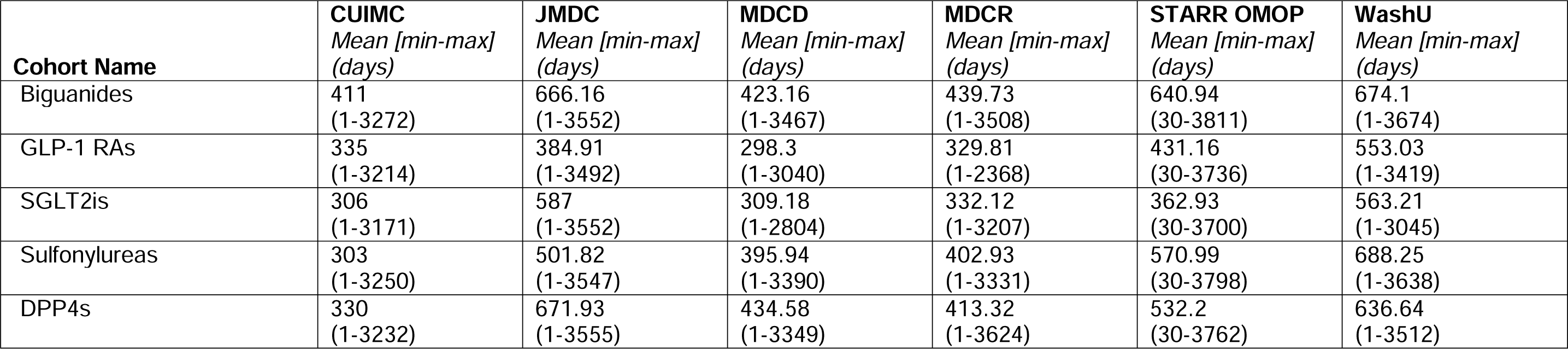
Drug Exposure Duration. Mean on-treatment follow-up time among adults with type 2 diabetes mellitus who met all study inclusion criteria, stratified by index glucose-lowering drug class and data source. Values are presented as the mean time with the minimum and maximum observed values shown in parentheses (mean [min–max]). A comprehensive list of time summaries for all drug classes, including cohorts explicitly excluding concomitant metformin use, is provided in the Supplementary Materials.

**Table 5.** Outcome Counts and Incidence Proportions. Outcome counts and incidence proportions (per 100 persons) among adults with type 2 diabetes mellitus who met the study inclusion criteria, stratified by data source. Values are presented as “*n* (incidence proportion per 100 persons)” over the follow-up period. Outcome-specific estimates stratified by drug exposure are reported in the Supplementary Materials.

| Outcome | CUIMC, *n* (%) | JMDC, *n* (%) | MDCD, *n* (%) | MDCR, *n* (%) | STARR OMOP, *n* (%) | WashU, *n* (%) |
| --- | --- | --- | --- | --- | --- | --- |
| Acute Pancreatitis | 476 (0.63) | 2141 (0.26) | 9015 (1.47) | 1268 (0.6) | 842 (0.99) | 1890 (1.31) |
| Hepatic Failure | 679 (0.97) | 1245 (0.16) | 6100 (1.09) | 1000 (0.49) | 1334 (1.73) | 1890 (1.46) |
| Hypoglycemia | 1004 (1.43) | 2189 (0.28) | 14496 (2.59) | 2113 (1.03) | 1107 (1.43) | 2680 (2.07) |
| Diabetic Ketoacidosis | 670 (0.9) | 1445 (0.18) | 6859 (1.11) | 323 (0.15) | 734 (0.87) | 1679 (1.16) |
| Abnormal Weight Gain | 492 (0.63) | 99 (0.01) | 20039 (3.3) | 2125 (1) | 1070 (1.25) | 0 (0) |
| Acute Renal Failure | 7084 (9.25) | 1624 (0.2) | 70021 (11.15) | 19123 (9.14) | 7614 (8.74) | 19628 (13.06) |
| Vomiting | 6544 (8.4) | 78072 (9.27) | 116193 (17.6) | 15249 (7.16) | 8788 (10.01) | 17161 (11.25) |
| Diarrhea | 6526 (8.23) | 7776 (0.94) | 91139 (13.95) | 19076 (8.78) | 7309 (8.14) | 16891 (11) |
| Stroke | 1745 (2.5) | 3319 (0.43) | 18031 (3.23) | 6533 (3.22) | 2469 (3.2) | 4347 (3.36) |
| Hospitalization with Heart Failure Events | 5888 (7.45) | 18974 (2.25) | 68741 (10.36) | 21068 (9.86) | 10210 (10.87) | 14840 (10.05) |
| Acute Myocardial Infarction | 2156 (2.96) | 2445 (0.3) | 23062 (3.82) | 7994 (3.82) | 4636 (5.46) | 4994 (3.59) |

Of our six databases, five (all but Washington University) contained subgroup comparisons with valid results that passed diagnostics.

For each subgroup, the top five pairwise drug comparisons with the lowest p-values were identified as potential signals of HTE (Table 6). Of these results, younger vs. older and younger vs. middle-aged subgroup comparisons failed study diagnostics. The renal impairment, poorly controlled diabetes, diabetic ketoacidosis, diabetic retinopathy, and Metabolic Dysfunction-Associated Steatotic Liver Disease (MASLD) subgroups also failed study diagnostics.

**Table 6.** Signals of Heterogeneity. Results for our HTE analysis. Hazard ratio estimates (HRs) and 95% confidence intervals from individual data sources were aggregated using random effects meta-analysis, followed by calibration using empirical null distributions to correct for residual confounding and systematic bias. The reported p-value corresponds to the statistical test of the difference between these subgroup-specific log hazard ratios, *Δln(HR)*. *cohort definition for drug exposure did not explicitly exclude metformin use **^†^**HR confidence interval does *not* include 1 and thus significantly favors one of the drug comparators Abbreviations: DPP-4i: dipeptidyl peptidase-4 inhibitor, GLP-1 RA: glucagon-like peptide-1 receptor agonist, HR: hazard ratio, SGLT-2i: sodium-glucose cotransporter 2 inhibitor, SU: sulfonylurea

| Hyperlipidemia (HLD) Subgroups (HLD vs. No HLD) |  |  |  |  |  |
| --- | --- | --- | --- | --- | --- |
| Outcome | Target | Comparator | HR (HLD) | HR (No HLD) | p-value |
| Abnormal weight gain | Biguanide | SU | 2.97 (0.92,9.56) | 0.45 (0.18,1.14) | **0.01** |
| Stroke | Biguanide | SGLT-2i | 1.76 (0.91,3.43) | 0.73 (0.44,1.23) | **0.04** |
| Acute myocardial infarction | Biguanide | DPP-4i | 0.55 (0.35,0.85)<sup>†</sup> | 1.13 (0.65,1.98) | **0.04** |
| Acute pancreatitis | Biguanide | DPP-4i | 0.74 (0.43,1.27) | 1.73 (0.89,3.36) | 0.05 |
| Diarrhea | Biguanide | DPP-4i | 0.97 (0.67,1.4) | 1.75 (1.08,2.82) | 0.06 |
| Hypertension (HTN) Subgroups (HTN vs. No HTN) |  |  |  |  |  |
| Outcome | Target | Comparator | HR (HTN) | HR (No HTN) | p-value |
| Acute myocardial infarction | Biguanide | SU | 1.65 (0.92, 2.95) | 0.78 (0.51,1.17) | **0.04** |
| Hospitalization with heart failure events | SU* | DPP-4i* | 0.84 (0.71, 1) | 1.07 (0.9, 1.28) | 0.05 |
| Hypoglycemia | Biguanide | DPP-4i | 0.89 (0.53, 1.5) | 0.45 (0.24, 0.84)<sub>†</sub> | 0.11 |
| Hypoglycemia | Biguanide | SGLT-2i | 1.37 (0.67, 2.79) | 0.54 (0.21,1.37) | 0.12 |
| Acute myocardial infarction | Biguanide | DPP-4i | 0.51 (0.27, 0.95)<sup>†</sup> | 0.9 (0.6, 1.35) | 0.14 |
| Obesity Subgroups (Obese vs. Non-Obese) |  |  |  |  |  |
| Outcome | Target | Comparator | HR (Obese) | HR (Non-Obese) | p-value |
| Hospitalization with heart failure events | Biguanide | DPP-4i | 0.53 (0.37, 0.75)<sup>†</sup> | 0.93 (0.76,1.15) | **0.01** |
| Stroke | SGLT-2i* | GLP-1 RA* | 1.5 (0.82, 2.72) | 0.51 (0.26, 0.99)<sub>†</sub> | **0.02** |
| Hypoglycemia | SU* | GLP-1 RA* | 1.21 (0.86, 1.7) | 2.35 (1.39, 3.97)<sub>†</sub> | **0.03** |
| Acute pancreatitis | Biguanide | SU | 1.96 (0.99, 3.88) | 0.82 (0.46, 1.47) | 0.06 |
| Hepatic failure | Biguanide | SU | 1.54 (0.73, 3.27) | 0.61 (0.32, 1.16) | 0.07 |
| Sex Subgroups (Male vs. Female) |  |  |  |  |  |
| Outcome | Target | Comparator | HR (Male) | HR (Female) | p-value |
| Diarrhea | GLP-1 RA* | DPP-4i* | 0.8 (0.49,1.28) | 1.43 (1.1, 1.84)<sup>†</sup> | **0.03** |
| Stroke | SGLT-2i* | DPP-4i* | 0.95 (0.68,1.32) | 0.38 (0.17, 0.86)<sub>†</sub> | **0.04** |
| Diarrhea | SU* | DPP-4i* | 0.84 (0.66,1.07) | 1.19 (0.95,1.5) | **0.04** |
| Diabetic ketoacidosis | GLP-1 RA* | DPP-4i* | 1.76 (0.77,4.05) | 0.66 (0.37,1.17) | 0.06 |
| Acute pancreatitis | SGLT-2i * | DPP-4i* | 1.28 (0.83,1.98) | 0.55 (0.21,1.42) | 0.11 |

**Table 6 Signals of Heterogeneity.**
| Age Subgroups (Older (>60 years) vs. Middle-aged (21-60 years)) |  |  |  |  |  |
| --- | --- | --- | --- | --- | --- |
| Outcome | Target | Comparator | HR (>60y) | HR (21-60y) | p-value |
| Diabetic ketoacidosis | Biguanide | GLP-1 RA | 0.73 (0.22, 2.4) | 2.84 (0.82, 9.8) | 0.12 |
| Hypoglycemia | SU* | GLP-1 RA* | 2.84 (1.51,5.33)<sup>†</sup> | 1.41 (0.74, 2.67) | 0.12 |
| Diabetic ketoacidosis | SU* | SGLT-2i * | 0.22 (0.06, 0.79) | 0.69 (0.34,1.37) | 0.12 |
| Abnormal weight gain | Biguanide | GLP-1 RA | 1.16 (0.44, 3.09) | 0.35 (0.11,1.17) | 0.13 |
| Acute renal failure | Biguanide | SGLT-2i | 2.15 (0.62,7.45) | 0.7 (0.29, 1.69) | 0.15 |

### Hyperlipidemia (HLD)

For acute myocardial infarctions, biguanides had a protective effect (compared to DPP-4i) in the HLD group, while it was not seen in the non-HLD group. While there is no definitive benefit or risk for taking biguanide vs. SU for abnormal weight gain, or for taking biguanide vs. SGLT-2i for stroke, our results show that there may exist a differential effect based on lipid status.

### Hypertension (HTN)

While there was no difference between risk of acute myocardial infarction between biguanide and SU, our results show that there may be a potential benefit of biguanides over SU only in the non-hypertensive subgroup.

*Obesity.* Biguanide had a protective effect (compared to DPP-4i) against hospitalization with heart failure only in the obese group, while SGLT-2i (compared to GLP-1 RA) had a protective effect against stroke only in the non-obese group. Finally, non-obese patients taking SU (vs. GLP-1 RA) had a higher risk of hypoglycemia, while the same effect was not seen for obese patients.

### Biological Sex

Female patients on GLP-1 RA (vs. DPP-4i) were more likely to experience diarrhea, and female patients on SGLT-2i (vs. DPP-4i) were more protected against stroke. These effects were not seen for male patients.

*Age*. None of the p-values were statistically significant in age-stratified analyses (Older (>60 years) vs. Middle-aged (21-60 years)), but the top 5 lowest p-values for HTE signal included subgroup differences for diabetic ketoacidosis (p = 0.12; biguanide vs. GLP-1 RA and SU vs. SGLT-2i) and hypoglycemia (p = 0.12, SU vs. GLP-1 RA).

## DISCUSSION

In this study, we identified several potential signals of HTE in demographic and clinical subgroups.

### Hyperlipidemia

Our results showed that there may be a benefit of biguanides to DPP-4i for protection against acute myocardial infarction only for those in the hyperlipidemia group. The literature reports DPP-4i to be protective against adverse cardiovascular events compared to metformin [35, 36], but studies did not explicitly adjust for dyslipidemia status.

While our results comparing biguanides and SGLT-2i are inconclusive in terms of benefit or risk against stroke (as individual HRs were not significantly different from 1), there was a difference in effect between hyperlipidemia and non-hyperlipidemia groups. Evidence in the literature is mixed: while some studies found that there was no difference in stroke risk [37] or risk in a composite cardiovascular outcome including stroke [38], one study found that SGLT-2i was *beneficial* for a composite cardiovascular outcome including stroke [39]. Furthermore, in a sub-analysis of the same study, where the patient cohort was restricted only to those with a diagnosis of *both* hypertension and hyperlipidemia, SGLT-2i was found to be beneficial compared to metformin [39]. Our results also suggest that stroke benefits of SGLT-2i may be further modified by hyperlipidemia status and warrants further investigation.

For abnormal weight gain, neither subgroup displayed a significant benefit or risk. However, it is well established that SU is *not* a weight-neutral treatment of diabetes [40–44], and our study suggests that, potentially, there is a differential effect of biguanide vs. SU depending on lipid status.

### Hypertension

Results in the literature of head-to-head comparisons of SU and metformin showed that patients taking SU had an *increased* cardiovascular event risk (acute myocardial infarction and stroke) even after stratifying by cardiovascular history [42], though SU did not affect cardiovascular mortality [41]. While these results did not explicitly stratify by blood pressure status, and the results from this current study did not show a significant benefit or harm from taking biguanide compared to SU, it is possible that there is a subgroup difference in the comparative effect of biguanide vs. SU.

### Obesity

Our results showed that the risk of hospitalization with heart failure events was *reduced* with biguanide (vs. DPP-4i) for obese patients, whereas that effect was not seen for non-obese patients. Previous studies have shown that metformin (vs. diet alone) reduced the risk of diabetes-related death and all-cause mortality in overweight and obese patients [45]. In contrast, the cardiovascular benefit of metformin in non-obese patients is less established: in a study where nonobese and obese patients were treated with metformin, the nonobese patients performed better regardless of the type of oral hypoglycemic agent used, because while the obese group lost more weight during the follow-up, the nonobese metformin group remained thinner [46]. Metformin is known to promote some weight loss, while DPP-4i is generally weight-neutral, thus a potential explanation for what we observed could be that because a higher BMI is associated with higher rates of heart failure events, the absolute benefit on metformin was larger in obese individuals.

For the outcome of stroke, SGLT-2i was more protective than GLP-1 RA for the non-obese subgroup, but not the obese subgroup. In the literature, both SGLT-2i and GLP-1RA have cardiovascular benefits and promote weight loss [40, 47–49], however there also exists potential subgroup heterogeneity: while the reduced risk of MACE conferred by GLP-1 RA persisted in both obese and non-obese subgroups, only obese subgroups benefitted from SGLT-2i for MACE risk [47, 48, 50]. Our results reflect this potential subgroup heterogeneity, though the direction of our heterogeneity conflicts with the literature, with *non-obese* subgroups benefiting from SGLT-2i vs. GLP-1 RA.

Finally, we observed a greater risk of hypoglycemia for non-obese patients taking SU (vs. GLP-1 RA), and this difference was not seen for obese patients. Hypoglycemia is a common side effect for patients receiving SU because of the drug’s mechanism of action (stimulating insulin release to lower blood glucose). In comparison, GLP-1 RA are thought to have a relatively low risk of hypoglycemia [40, 51]. A head-to-head comparison of liraglutide and glimepiride, in conjunction with metformin, showed that the incidence of hypoglycemia on liraglutide was lower than that of glimepiride [52–54]. Thus, it is not surprising that the risk of hypoglycemia was increased on SU vs. GLP-1 RA. Furthermore, our finding here potentially reflects the inverse relationship between BMI and hypoglycemia [55, 56].

### Biological Sex

Some of the most common GLP-1 RA side effects are gastrointestinal (ex, vomiting, nausea, diarrhea) [57–59]. Our results additionally showed a differential risk of female patients experiencing diarrhea more often on GLP-1 RA (vs. DPP-4i), which is consistent with literature showing that women experience GI side effects with GLP-1 RAs at roughly twice the rate of men [60–62]. Similarly, our finding that the protective effect of SGLT-2i (compared to DPP-4i) for female patients was greater than what was seen for male patients is consistent with findings in the literature that in general, risk reductions for cardiovascular outcomes is higher in women vs. men [63]. Potentially, the differences we observed with diarrhea and stroke risk can be attributed to gender differences in adverse event reporting [64] or biological differences in cardiovascular disease risk factors [65, 66].

### Age

None of the subgroup comparisons were significantly different in our study. However, we did see a significant increase in risk of hypoglycemia older patients (>60 years) on SU vs GLP-1 RA, even if the difference in the risk was *not* significantly different between older and younger (21-60 years) patients. As previously discussed, this reflects the known risk of hypoglycemia for patients taking SU [40]. This finding supports current guidelines recommending caution with sulfonylureas in older adults, given their susceptibility to hypoglycemia-related adverse events [67], which can subsequently increase risks of falls and hospitalizations [68, 69].

### Limitations

There remain limitations to our study. First, as with any observational analysis, residual confounding may be a concern. Although we mitigated this through propensity score matching, verified covariate balance using standardized mean differences, and calibrated our effect estimates with negative control outcomes, unmeasured confounding cannot be excluded. As such, causal conclusions regarding the relationship between drug exposures and adverse events should be drawn with caution.

Second, the available sample size may have limited the detection of HTE. Stratification by subgroups inherently reduces the number of patients available for each comparison, which may obscure weaker signals of effect modification. In addition, many of the reported comparisons arose in single databases, limiting generalizability. Where results from multiple databases were available, we included all estimates in the meta-analysis, but replication across patient populations would further strengthen confidence in our findings. These limitations may be partially addressed by refinement of phenotype and subgroup definitions (e.g., categorizing renal impairment as two subgroups rather than three) and recruiting additional data partners to improve the generalizability of our findings.

## Data Availability Statement

The detailed study protocol, including concept sets for our cohorts, and the analysis code can be found in https://github.com/ohdsi-studies/LegendT2dmArpah.

## Author’s relationships and activities

AO is an employee of Johnson & Johnson. GH and MAS have received grant funding from Johnson & Johnson to support methods research not directly related to this study. Johnson & Johnson did not have input in the design, execution, interpretation of results or decision to publish. MAS has contracts and grants from Johnson & Johnson, Gilead and the US Food & Drug Administration.

All other authors have no competing interests.

## Contribution statement

All authors reviewed and edited the manuscript for accuracy, intellectual content, clarity, and coherence, and approved the final version for submission. In addition:

HYC contributed to the study design, statistical analysis plan, and interpretation of the findings. She drafted the initial manuscript.

TF contributed to the study design, statistical analysis plan, execution of the analyses, coordinated data partner communications, and interpretation of the findings.

TVA, XJ, and DDG contributed to the study design, statistical analysis plan, and interpretation of the findings.

AO, LZ, RF, and HMC executed the analyses at their respective data sites and contributed to data quality review.

MAS and GH supervised the study, contributed to its conceptualization and design, and provided critical input on data interpretation.

## Abbreviations

BMI: Body Mass Index
CDM: Common Data Model
DKA: Diabetic Ketoacidosis
DPP-4i: Dipeptidyl Peptidase-4 Inhibitors
EHR: Electronic Health Records
EASE: Expected Absolute Systematic Error
GLP-1 RA: Glucagon-Like Peptide-1 Receptor Agonists
HbA1c: Hemoglobin A1c
HLD: Hyperlipidemia
HR: Hazard Ratio
HTE: Heterogeneity of Treatment Effect
HTN: Hypertension
LEGEND-HTN: Large-scale Evidence Generation and Evaluation: Hypertension
LEGEND-T2DM: Large-scale Evidence Generation and Evaluation: Type 2 Diabetes Mellitus
MACE: Major Adverse Cardiac Events
MASLD: Metabolic Dysfunction-Associated Steatotic Liver Disease
MDRR: Minimum Detectable Relative Risk
OHDSI: Observational Health Data Sciences and Informatics
OMOP: Observational Medical Outcomes Partnership
RCT: Randomized Controlled Trials
SGLT2-i: Sodium-Glucose Co-Transporter-2 Inhibitors
SU: Sulfonylureas

## REFERENCES

1. 2024 Heart Disease and Stroke Statistics: A Report of US and Global Data From the American Heart Association. https://www.ahajournals.org/doi/epub/10.1161/CIR.0000000000001209. Accessed 14 May 2025

2. Beckman JA, Creager MA, Libby P (2002) Diabetes and AtherosclerosisEpidemiology, Pathophysiology, and Management. JAMA 287(19):2570–2581. 10.1001/jama.287.19.2570

3. Thomas MC, Cooper ME, Zimmet P (2016) Changing epidemiology of type 2 diabetes mellitus and associated chronic kidney disease. Nat Rev Nephrol 12(2):73–81. 10.1038/nrneph.2015.173

4. Zhang L, Li X, Wolfe CDA, O’Connell MDL, Wang Y (2021) Diabetes As an Independent Risk Factor for Stroke Recurrence in Ischemic Stroke Patients: An Updated Meta-Analysis. Neuroepidemiology 55(6):427–435. 10.1159/000519327

5. Gregg EW, Pratt A, Owens A, et al (2024) The burden of diabetes-associated multiple long-term conditions on years of life spent and lost. Nat Med 30(10):2830–2837. 10.1038/s41591-024-03123-2

6. Gregg EW, Li Y, Wang J, et al (2014) Changes in Diabetes-Related Complications in the United States, 1990–2010. New England Journal of Medicine 370(16):1514–1523. 10.1056/NEJMoa1310799

7. Nathan DM (1993) Long-Term Complications of Diabetes Mellitus. New England Journal of Medicine 328(23):1676–1685. 10.1056/NEJM199306103282306

8. Solomon SD, McMurray JJV, Claggett B, et al (2022) Dapagliflozin in Heart Failure with Mildly Reduced or Preserved Ejection Fraction. N Engl J Med 387(12):1089–1098. 10.1056/NEJMoa2206286

9. Hernandez AF, Green JB, Janmohamed S, et al (2018) Albiglutide and cardiovascular outcomes in patients with type 2 diabetes and cardiovascular disease (Harmony Outcomes): a double-blind, randomised placebo-controlled trial. Lancet 392(10157):1519–1529. 10.1016/S0140-6736(18)32261-X

10. Andrikou E, Tsioufis C, Andrikou I, Leontsinis I, Tousoulis D, Papanas N (2019) GLP-1 receptor agonists and cardiovascular outcome trials: An update. Hellenic J Cardiol 60(6):347–351. 10.1016/j.hjc.2018.11.008

11. Pablo A, Evelyn B, Claudia F, Yanina MA (2021) GLP-1RA and SGLT2i: Cardiovascular Impact on Diabetic Patients. Curr Hypertens Rev 17(2):149–158. 10.2174/1573402116999201124123549

12. Khera R, Schuemie MJ, Lu Y, et al (2022) Large-scale evidence generation and evaluation across a network of databases for type 2 diabetes mellitus (LEGEND-T2DM): a protocol for a series of multinational, real-world comparative cardiovascular effectiveness and safety studies. BMJ Open 12(6):e057977. 10.1136/bmjopen-2021-057977

13. Rohan Khera MD, Arya Aminorroaya MD, Lovedeep Singh Dhingra M, et al (2024) Comparative Effectiveness of Second-Line Antihyperglycemic Agents for Cardiovascular Outcomes: A Multinational, Federated Analysis of LEGEND-T2DM. Journal of the American College of Cardiology. 10.1016/j.jacc.2024.05.069

14. Trischitta V, Prudente S, Doria A (2020) Disentangling the heterogeneity of adulthood-onset non-autoimmune diabetes: a little closer but lot more to do. Current Opinion in Pharmacology 55:157–164. 10.1016/j.coph.2020.10.020

15. Ahlqvist E, Storm P, Käräjämäki A, et al (2018) Novel subgroups of adult-onset diabetes and their association with outcomes: a data-driven cluster analysis of six variables. The Lancet Diabetes & Endocrinology 6(5):361–369. 10.1016/S2213-8587(18)30051-2

16. Nair ATN, Wesolowska-Andersen A, Brorsson C, et al (2022) Heterogeneity in phenotype, disease progression and drug response in type 2 diabetes. Nat Med 28(5):982–988. 10.1038/s41591-022-01790-7

17. Redondo MJ, Hagopian WA, Oram R, et al (2020) The clinical consequences of heterogeneity within and between different diabetes types. Diabetologia 63(10):2040– 2048. 10.1007/s00125-020-05211-7

18. Gloyn AL, Drucker DJ (2018) Precision medicine in the management of type 2 diabetes. The Lancet Diabetes & Endocrinology 6(11):891–900. 10.1016/S2213-8587(18)30052-4

19. Florez JC (2017) Pharmacogenetics in type 2 diabetes: precision medicine or discovery tool? Diabetologia 60(5):800–807. 10.1007/s00125-017-4227-1

20. Dávila-García DM, Falconer T, Pratt N, Natarajan K, Hripcsak G (2025) Heterogeneity of treatment effects of glucose-lowering drug classes for type 2 diabetes: LEGEND-T2DM network real-world evidence. Journal of Diabetes and its Complications 39(9):109114. 10.1016/j.jdiacomp.2025.109114

21. Suchard MA, Schuemie MJ, Krumholz HM, et al (2019) Comprehensive comparative effectiveness and safety of first-line antihypertensive drug classes: a systematic, multinational, large-scale analysis. The Lancet 394(10211):1816–1826. 10.1016/S0140-6736(19)32317-7

22. (2024) OMOP Common Data Model

23. Rao G (2025) PhenotypeLibrary: The OHDSI Phenotype Library

24. Tian Y, Schuemie MJ, Suchard MA (2018) Evaluating large-scale propensity score performance through real-world and synthetic data experiments. Int J Epidemiol 47(6):2005–2014. 10.1093/ije/dyy120

25. Zhang L, Wang Y, Schuemie MJ, Blei DM, Hripcsak G (2022) Adjusting for indirectly measured confounding using large-scale propensity score. Journal of Biomedical Informatics 134:104204. 10.1016/j.jbi.2022.104204

26. Schuemie MJ, Hripcsak G, Ryan PB, Madigan D, Suchard MA (2016) Robust empirical calibration of p-values using observational data. Stat Med 35(22):3883–3888. 10.1002/sim.6977

27. Suchard MA, Simpson SE, Zorych I, Ryan P, Madigan D (2013) Massive parallelization of serial inference algorithms for a complex generalized linear model. ACM Trans Model Comput Simul 23(1). 10.1145/2414416.2414791

28. Schuemie M, Reps J, Black A, et al (2024) Health-Analytics Data to Evidence Suite (HADES): Open-Source Software for Observational Research. Stud Health Technol Inform 310:966–970. 10.3233/SHTI231108

29. Sena A, Schuemie M, Gilbert J (2025) Strategus: Coordinate and Execute OHDSI HADES Modules.

30. Viechtbauer W (2009) metafor: Meta-Analysis Package for R. 4.6–0

31. Conover MM, Ryan PB, Chen Y, Suchard MA, Hripcsak G, Schuemie MJ (2025) Objective study validity diagnostics: a framework requiring pre-specified, empirical verification to increase trust in the reliability of real-world evidence. J Am Med Inform Assoc 32(3):518–525. 10.1093/jamia/ocae317

32. Brody T (2012) Chapter 9 - Biostatistics. In: Brody T (ed) Clinical Trials. Academic Press, Boston, pp 165–190

33. Bewick V, Cheek L, Ball J (2004) Statistics review 12: Survival analysis. Critical Care 8(5):389. 10.1186/cc2955

34. Hedges LV, Vevea JL (1998) Fixed- and random-effects models in meta-analysis. Psychological Methods 3(4):486–504. 10.1037/1082-989X.3.4.486

35. Noguchi Y, Yoshizawa S, Tachi T, Teramachi H (2022) Effect of Dipeptidyl Peptidase-4 Inhibitors vs. Metformin on Major Cardiovascular Events Using Spontaneous Reporting System and Real-World Database Study. J Clin Med 11(17):4988. 10.3390/jcm11174988

36. Wu D, Li L, Liu C (2014) Efficacy and safety of dipeptidyl peptidase-4 inhibitors and metformin as initial combination therapy and as monotherapy in patients with type 2 diabetes mellitus: a meta-analysis. Diabetes Obes Metab 16(1):30–37. 10.1111/dom.12174

37. Chen T-H, Li Y-R, Chen S-W, et al (2020) Sodium-glucose cotransporter 2 inhibitor versus metformin as first-line therapy in patients with type 2 diabetes mellitus: a multi-institution database study. Cardiovascular Diabetology 19(1):189. 10.1186/s12933-020-01169-3

38. Shin H, Schneeweiss S, Glynn RJ, Patorno E (2022) Cardiovascular Outcomes in Patients Initiating First-Line Treatment of Type 2 Diabetes With Sodium–Glucose Cotransporter-2 Inhibitors Versus Metformin. Ann Intern Med 175(7):927–937. 10.7326/M21-4012

39. Fralick M, Schneeweiss S, Redelmeier DA, Razak F, Gomes T, Patorno E (2021) Comparative effectiveness and safety of sodium-glucose cotransporter-2 inhibitors versus metformin in patients with type 2 diabetes: An observational study using data from routine care. Diabetes, Obesity and Metabolism 23(10):2320–2328. 10.1111/dom.14474

40. Apovian CM, Okemah J, O’Neil PM (2019) Body Weight Considerations in the Management of Type 2 Diabetes. Adv Ther 36(1):44–58. 10.1007/s12325-018-0824-8

41. Hemmingsen B, Schroll JB, Wetterslev J, et al (2014) Sulfonylurea versus metformin monotherapy in patients with type 2 diabetes: a Cochrane systematic review and meta-analysis of randomized clinical trials and trial sequential analysis. CMAJ Open 2(3):E162–E175. 10.9778/cmajo.20130073

42. Roumie CL, Hung AM, Greevy RA, et al (2012) Comparative Effectiveness of Sulfonylurea and Metformin Monotherapy on Cardiovascular Events in Type 2 Diabetes Mellitus. Ann Intern Med 157(9):601–610. 10.7326/0003-4819-157-9-201211060-00003

43. Kahn SE, Haffner SM, Heise MA, et al (2006) Glycemic Durability of Rosiglitazone, Metformin, or Glyburide Monotherapy. New England Journal of Medicine 355(23):2427–2443. 10.1056/NEJMoa066224

44. Campbell IW, Menzies DG, Chalmers J, McBain AM, Brown IR (1994) One year comparative trial of metformin and glipizide in type 2 diabetes mellitus. Diabete Metab 20(4):394–400

45. UKPDS Group (1998) Effect of intensive blood-glucose control with metformin on complications in overweight patients with type 2 diabetes (UKPDS 34). The Lancet 352(9131):854–865. 10.1016/S0140-6736(98)07037-8

46. Ong CR, Molyneaux LM, Constantino MI, Twigg SM, Yue DK (2006) Long-Term Efficacy of Metformin Therapy in Nonobese Individuals With Type 2 Diabetes. Diabetes Care 29(11):2361–2364. 10.2337/dc06-0827

47. Diallo A, Carlos-Bolumbu M, Galtier F (2022) Age, sex, race, BMI, and duration of diabetes differences in cardiovascular outcomes with glucose lowering drugs in type 2 diabetes: A systematic review and meta-analysis. eClinicalMedicine 54. 10.1016/j.eclinm.2022.101697

48. Sattar N, Lee MMY, Kristensen SL, et al (2021) Cardiovascular, mortality, and kidney outcomes with GLP-1 receptor agonists in patients with type 2 diabetes: a systematic review and meta-analysis of randomised trials. The Lancet Diabetes & Endocrinology 9(10):653–662. 10.1016/S2213-8587(21)00203-5

49. Goldenberg RM, Cheng AYY, Fitzpatrick T, Gilbert JD, Verma S, Hopyan JJ (2022) Benefits of GLP-1 (Glucagon-Like Peptide 1) Receptor Agonists for Stroke Reduction in Type 2 Diabetes: A Call to Action for Neurologists. Stroke 53(5):1813–1822. 10.1161/STROKEAHA.121.038151

50. Kristensen SL, Rørth R, Jhund PS, et al (2019) Cardiovascular, mortality, and kidney outcomes with GLP-1 receptor agonists in patients with type 2 diabetes: a systematic review and meta-analysis of cardiovascular outcome trials. The Lancet Diabetes & Endocrinology 7(10):776–785. 10.1016/S2213-8587(19)30249-9

51. Nauck M (2016) Incretin therapies: highlighting common features and differences in the modes of action of glucagon-like peptide-1 receptor agonists and dipeptidyl peptidase-4 inhibitors. Diabetes, Obesity and Metabolism 18(3):203–216. 10.1111/dom.12591

52. Nauck M, Frid A, Hermansen K, et al (2009) Efficacy and Safety Comparison of Liraglutide, Glimepiride, and Placebo, All in Combination With Metformin, in Type 2 Diabetes. Diabetes Care 32(1):84–90. 10.2337/dc08-1355

53. Shyangdan DS, Royle P, Clar C, Sharma P, Waugh N, Snaith A (2011) Glucagon-like peptide analogues for type 2 diabetes mellitus. Cochrane Database of Systematic Reviews. 10.1002/14651858.cd006423.pub2

54. Yang W, Chen L, Ji Q, et al (2011) Liraglutide provides similar glycaemic control as glimepiride (both in combination with metformin) and reduces body weight and systolic blood pressure in Asian population with type 2 diabetes from China, South Korea and India: a 16-week, randomized, double-blind, active control trial. Diabetes, Obesity and Metabolism 13(1):81–88. 10.1111/j.1463-1326.2010.01323.x

55. Yun J-S, Park Y-M, Han K, Cha S-A, Ahn Y-B, Ko S-H (2019) The Association Between BMI and the Risk of Severe Hypoglycemia in Type 2 Diabetes. Diabetes Metab 45(1):19–25. 10.1016/j.diabet.2018.03.006

56. Plečko D, Bennett N, Mårtensson J, Bellomo R (2021) The obesity paradox and hypoglycemia in critically ill patients. Critical Care 25(1):378. 10.1186/s13054-021-03795-z

57. Nauck MA, Meier JJ (2019) Are all GLP-1 agonists equal in the treatment of type 2 diabetes? Eur J Endocrinol 181(6):R211–R234. 10.1530/EJE-19-0566

58. Nauck MA, Quast DR, Wefers J, Meier JJ (2021) GLP-1 receptor agonists in the treatment of type 2 diabetes – state-of-the-art. Molecular Metabolism 46:101102. 10.1016/j.molmet.2020.101102

59. Bettge K, Kahle M, Abd El Aziz MS, Meier JJ, Nauck MA (2017) Occurrence of nausea, vomiting and diarrhoea reported as adverse events in clinical trials studying glucagon-like peptide-1 receptor agonists: A systematic analysis of published clinical trials. Diabetes, Obesity and Metabolism 19(3):336–347. 10.1111/dom.12824

60. Joung K-I, Jung G-W, Park H-H, Lee H, Park S-H, Shin J-Y (2020) Gender differences in adverse event reports associated with antidiabetic drugs. Sci Rep 10(1):17545. 10.1038/s41598-020-74000-4

61. de Vries ST, Denig P, Ekhart C, Mol PGM, van Puijenbroek EP (2020) Sex Differences in Adverse Drug Reactions of Metformin: A Longitudinal Survey Study. Drug Saf 43(5):489–495. 10.1007/s40264-020-00913-8

62. Roseberry T, Grossrubatscher I, Krausz T, Wang Y, Schwartz M, Tingley D (2025) Sex differences in GLP-1 signaling across species. 2025.03.17.643822

63. Raparelli V, Elharram M, Moura CS, et al (2020) Sex Differences in Cardiovascular Effectiveness of Newer Glucose-Lowering Drugs Added to Metformin in Type 2 Diabetes Mellitus. Journal of the American Heart Association 9(1):e012940. 10.1161/JAHA.119.012940

64. Watson S, Caster O, Rochon PA, den Ruijter H (2019) Reported adverse drug reactions in women and men: Aggregated evidence from globally collected individual case reports during half a century. EClinicalMedicine 17:100188. 10.1016/j.eclinm.2019.10.001

65. Arnetz L, Ekberg NR, Alvarsson M (2014) Sex differences in type 2 diabetes: focus on disease course and outcomes. Diabetes Metab Syndr Obes 7:409–420. 10.2147/DMSO.S51301

66. Mauvais-Jarvis F (2018) Gender differences in glucose homeostasis and diabetes. Physiology & Behavior 187:20–23. 10.1016/j.physbeh.2017.08.016

67. Abdelhafiz AH, Rodríguez-Mañas L, Morley JE, Sinclair AJ (2015) Hypoglycemia in Older People - A Less Well Recognized Risk Factor for Frailty. Aging Dis 6(2):156–167. 10.14336/AD.2014.0330

68. (2022) 13. Older Adults: Standards of Medical Care in Diabetes—2022. Diabetes Care 45(Suppl 1):S195–S207. 10.2337/dc22-S013

69. Ling S, Zaccardi F, Lawson C, Seidu SI, Davies MJ, Khunti K (2021) Glucose Control, Sulfonylureas, and Insulin Treatment in Elderly People With Type 2 Diabetes and Risk of Severe Hypoglycemia and Death: An Observational Study. Diabetes Care 44(4):915–924. 10.2337/dc20-0876

