## Supplementary Materials for "Heterogeneity of Treatment Effects Across Nine Glucose-Lowering Drug Classes in Type 2 Diabetes: Extension of the LEGEND-T2DM Network Study"

### Supplementary Tables

| Data source | Description |
| --- | --- |
| Columbia University Irving Medical Center (CUIMC) | The clinical data warehouse of NewYork-Presbyterian Hospital/Columbia University Irving Medical Center, New York, NY, based on its current and previous electronic health record systems, with data spanning over 30 years and including over 6 million patients |
| Merative MarketScan Multi-State Medicaid Database (MDCD) | Merative MarketScan Multi-State Medicaid Database (MDCD) contains adjudicated US health insurance claims for Medicaid enrollees from multiple states and includes hospital discharge diagnoses, outpatient diagnoses and procedures, and outpatient pharmacy claims as well as ethnicity and Medicare eligibility. Members maintain their same identifier even if they leave the system for a brief period; however the dataset lacks lab data. |
| Merative MarketScan Medicare Supplemental and Coordination of Benefits Database (MDCR) | Merative MarketScan Medicare Supplemental and Coordination of Benefits Database (MDCR) represents health services of retirees in the United States with primary or Medicare supplemental coverage through privately insured fee-for-service, point-of-service, or capitated health plans. These data include adjudicated health insurance claims (e.g. inpatient, outpatient, and outpatient pharmacy).Additionally, it captures laboratory tests for a subset of the covered lives. |
| STARR-OMOP (Stanford Healthcare) | STARR-OMOP is the clinical data warehouse of Stanford Medicine, containing de-identified patient data from Stanford Health Care and Stanford Children's Health electronic health record (EHR) systems. It contains data for more than 3.7 million patients. |
| JMDC Claims Database | The JMDC Claims Database is an epidemiological receipt database that has accumulated receipts (inpatient, outpatient, dispensing) and medical examination data. |
| Washington University in St. Louis (WashU) | Clinical electronic health record for Washington University in St. Louis, a non-profit academic medical center |

**Supplementary Table S1 Database Descriptions.** Descriptions of the six databases in the study.

| Cohort ID | Cohort Name | Outcome Concept ID |
|---|---|---|
| 101 | Allergic rhinitis | 257007 |
| 102 | Carpal tunnel syndrome | 380094 |
| 103 | Cerebral palsy | 4134120 |
| 104 | Chronic obstructive lung disease | 255573 |
| 105 | Contact dermatitis | 134438 |
| 106 | Cyst of ovary | 197610 |
| 107 | Deviated nasal septum | 377910 |
| 108 | Dislocation of shoulder joint | 4213373 |
| 109 | Endometriosis (clinical) | 433527 |
| 110 | Foreign body in ear | 374801 |
| 111 | Gout | 440674 |
| 112 | Hemorrhoids | 195562 |
| 113 | Hypoparathyroidism | 140362 |
| 114 | Influenza | 4266367 |
| 115 | Ingrowing nail | 139099 |
| 116 | Osteoarthritis of knee | 4079750 |
| 117 | Prostatitis | 194997 |
| 118 | Sciatica | 372409 |
| 119 | Sleep apnea | 313459 |
| 120 | Vitamin D deficiency | 436070 |

**Supplementary Table S2 List of Negative Control Outcomes.** For each target-comparator comparison and outcome within each database, we conducted negative control outcome experiments where the null hypothesis of no effect is believed to be true, using this set of 20 control outcomes.

| Cohort Name | CUIMC, *n* | JMDC, *n* | MDCD, *n* | MDCR, *n* | STARR OMOP, *n* | WashU, *n* |
|---|---|---|---|---|---|---|
| **All T2DM Patients** | **138367** | **854819** | **873887** | **756418** | **111674** | **296727** |
| **T2DM patients fulfilling inclusion criteria** | **70189** | **782022** | **561171** | **205698** | **77421** | **129379** |
| **SGLT2is excluding concomitant metformin use** | **2302** | **43573** | **4695** | **4826** | **3098** | **677** |
| Male | 1409 | 32005 | 2513 | 2769 | 1874 | 418 |
| Female | 893 | 11568 | 2182 | 2057 | 1224 | 259 |
| Age <=20 | -5 | 144 | 17 | NA | 10 | NA |
| Age 21-60 | 711 | 34188 | 3101 | 12 | 940 | 270 |
| Age 61-80 | 1268 | 9241 | 1469 | 3345 | 1750 | 346 |
| HbA1c > 8% | 66 | NA | NA | NA | 51 | -5 |
| History of MASLD | 14 | NA | 56 | 20 | 35 | -5 |
| Renal Impairment not on Dialysis | 824 | 10454 | 2157 | 2249 | 1043 | 185 |
| Renal Impairment on Dialysis | 42 | 311 | 112 | 44 | 104 | 6 |
| Obese | 1286 | 15523 | 2559 | 1759 | 1260 | 327 |
| History of DKA | 15 | 309 | 84 | 19 | 19 | -5 |
| History of Diabetic Retinopathy | 59 | NA | 92 | 36 | 66 | 7 |
| History of Hyperlipidemia | 1466 | 29483 | 3025 | 2481 | 1614 | 401 |
| History of Hypertension | 1585 | 27964 | 3419 | 2455 | 1633 | 405 |
| **Sulfonylureas excluding concomitant metformin use** | **2593** | **2867** | **7812** | **3274** | **4654** | **928** |
| Male | 1328 | 2079 | 2912 | 1839 | 2580 | 524 |
| Female | 1264 | 788 | 4900 | 1435 | 2072 | 404 |
| Age <=20 | -5 | 23 | 137 | NA | 11 | -5 |
| Age 21-60 | 688 | 2275 | 5829 | 27 | 1526 | 275 |
| Age 61-80 | 1388 | 569 | 1566 | 2284 | 2526 | 503 |
| HbA1c > 8% | 86 | NA | NA | NA | 129 | 6 |
| History of MASLD | -5 | NA | 71 | 11 | 42 | -5 |
| Renal Impairment not on Dialysis | 503 | 377 | 2001 | 1463 | 972 | 163 |
| Renal Impairment on Dialysis | 62 | 6 | 234 | 81 | 301 | 14 |
| Obese | 1040 | 810 | 3587 | 1007 | 1405 | 330 |
| History of DKA | 19 | 32 | 164 | 31 | 22 | 5 |
| History of Diabetic Retinopathy | 29 | NA | 162 | 64 | 110 | -5 |
| History of Hyperlipidemia | 1104 | 1561 | 3602 | 2345 | 1811 | 433 |
| History of Hypertension | 1419 | 1342 | 4563 | 2571 | 2091 | 515 |
| **DPP4s excluding concomitant metformin use** | **2230** | **47436** | **2211** | **1744** | **1644** | **310** |
| Male | 1111 | 34090 | 895 | 878 | 800 | 161 |
| Female | 1118 | 13346 | 1316 | 866 | 844 | 149 |
| Age <=20 | NA | 112 | 7 | NA | -5 | NA |
| Age 21-60 | 523 | 35605 | 1292 | 22 | 402 | 91 |
| Age 61-80 | 1193 | 11719 | 713 | 1141 | 950 | 161 |
| HbA1c > 8% | 100 | NA | NA | NA | 61 | -5 |
| History of MASLD | -5 | NA | 25 | 9 | 16 | -5 |
| Renal Impairment not on Dialysis | 600 | 6657 | 792 | 855 | 354 | 83 |

| | | | | | | |
|---|---|---|---|---|---|---|
| Renal Impairment on Dialysis | 99 | 261 | 155 | 54 | 98 | 5 |
| Obese | 860 | 11896 | 967 | 522 | 473 | 109 |
| History of DKA | 30 | 342 | 63 | 30 | 8 | -5 |
| History of Diabetic Retinopathy | 47 | NA | 106 | 41 | 51 | -5 |
| History of Hyperlipidemia | 1062 | 29582 | 1330 | 1290 | 650 | 158 |
| History of Hypertension | 1353 | 25398 | 1557 | 1390 | 731 | 188 |
| **Alpha-glucosidase inhibitors** | **-5** | **1752** | **5** | **-5** | **-5** | **NA** |
| Male | -5 | 1154 | NA | -5 | -5 | NA |
| Female | NA | 598 | 5 | -5 | NA | NA |
| Age <=20 | NA | 24 | NA | NA | NA | NA |
| Age 21-60 | -5 | 1394 | -5 | NA | NA | NA |
| Age 61-80 | NA | 334 | -5 | -5 | -5 | NA |
| HbA1c > 8% | -5 | NA | NA | NA | NA | NA |
| Renal Impairment not on Dialysis | -5 | 315 | -5 | -5 | NA | NA |
| Renal Impairment on Dialysis | NA | 9 | NA | NA | NA | NA |
| Obese | -5 | 399 | -5 | -5 | NA | NA |
| History of DKA | NA | 23 | NA | NA | NA | NA |
| History of Hyperlipidemia | -5 | 1017 | -5 | -5 | NA | NA |
| History of Hypertension | -5 | 760 | -5 | -5 | NA | NA |
| **Alpha-glucosidase inhibitors excluding concomitant metformin use** | **-5** | **1340** | **-5** | **-5** | **-5** | **NA** |
| Male | -5 | 881 | NA | -5 | -5 | NA |
| Female | NA | 459 | -5 | NA | NA | NA |
| Age <=20 | NA | 15 | NA | NA | NA | NA |
| Age 21-60 | -5 | 1039 | -5 | NA | NA | NA |
| Age 61-80 | NA | 286 | -5 | -5 | -5 | NA |
| Renal Impairment not on Dialysis | NA | 227 | -5 | -5 | NA | NA |
| Renal Impairment on Dialysis | NA | 8 | NA | NA | NA | NA |
| Obese | NA | 250 | -5 | NA | NA | NA |
| History of DKA | NA | 14 | NA | NA | NA | NA |
| History of Hyperlipidemia | NA | 765 | -5 | -5 | NA | NA |
| History of Hypertension | NA | 574 | -5 | -5 | NA | NA |
| **GIP-GLP-1 RA dual agonists** | **180** | **NA** | **554** | **963** | **759** | **302** |
| Male | 70 | NA | 114 | 363 | 301 | 146 |
| Female | 110 | NA | 440 | 600 | 458 | 156 |
| Age <=20 | NA | NA | 6 | NA | -5 | NA |
| Age 21-60 | 107 | NA | 469 | 15 | 446 | 226 |
| Age 61-80 | 69 | NA | 79 | 914 | 296 | 75 |
| HbA1c > 8% | -5 | NA | NA | NA | 17 | NA |
| History of MASLD | -5 | NA | 11 | 17 | 26 | 6 |
| Renal Impairment not on Dialysis | 22 | NA | 113 | 267 | 139 | 26 |
| Renal Impairment on Dialysis | -5 | NA | -5 | -5 | 5 | -5 |

| | | | | | | |
|---|---|---|---|---|---|---|
| Obese | 165 | NA | 426 | 659 | 636 | 260 |
| History of DKA | -5 | NA | 10 | 5 | 8 | -5 |
| History of Diabetic Retinopathy | -5 | NA | 8 | -5 | 14 | NA |
| History of Hyperlipidemia | 135 | NA | 331 | 661 | 512 | 158 |
| History of Hypertension | 119 | NA | 335 | 657 | 486 | 167 |
| **GIP-GLP-1 RA dual agonists excluding concomitant metformin use** | **81** | **NA** | **276** | **678** | **341** | **155** |
| Male | 32 | NA | 58 | 238 | 121 | 62 |
| Female | 49 | NA | 218 | 440 | 220 | 93 |
| Age <=20 | NA | NA | -5 | NA | NA | NA |
| Age 21-60 | 49 | NA | 230 | 11 | 188 | 110 |
| Age 61-80 | 31 | NA | 42 | 640 | 144 | 44 |
| HbA1c > 8% | -5 | NA | NA | NA | 7 | NA |
| History of MASLD | -5 | NA | 7 | 10 | 9 | -5 |
| Renal Impairment not on Dialysis | 13 | NA | 55 | 188 | 61 | 15 |
| Renal Impairment on Dialysis | -5 | NA | NA | -5 | -5 | -5 |
| Obese | 75 | NA | 206 | 466 | 290 | 122 |
| History of DKA | -5 | NA | -5 | NA | -5 | -5 |
| History of Diabetic Retinopathy | -5 | NA | 6 | NA | 5 | NA |
| History of Hyperlipidemia | 62 | NA | 161 | 465 | 226 | 69 |
| History of Hypertension | 52 | NA | 162 | 462 | 224 | 84 |
| **Thiazolidinediones** | **583** | **2301** | **2514** | **893** | **1136** | **241** |
| Male | 292 | 1700 | 1016 | 503 | 665 | 146 |
| Female | 291 | 601 | 1498 | 390 | 471 | 95 |
| Age <=20 | -5 | 13 | 119 | NA | -5 | NA |
| Age 21-60 | 151 | 1963 | 1929 | 8 | 324 | 85 |
| Age 61-80 | 343 | 325 | 436 | 709 | 668 | 126 |
| HbA1c > 8% | 31 | NA | NA | NA | 31 | -5 |
| History of MASLD | -5 | NA | 39 | 11 | 8 | NA |
| Renal Impairment not on Dialysis | 100 | 316 | 536 | 308 | 188 | 39 |
| Renal Impairment on Dialysis | -5 | -5 | 24 | 9 | 14 | -5 |
| Obese | 230 | 776 | 1440 | 307 | 376 | 100 |
| History of DKA | -5 | 17 | 86 | 8 | 7 | -5 |
| History of Diabetic Retinopathy | -5 | NA | 82 | 36 | 33 | -5 |
| History of Hyperlipidemia | 294 | 1628 | 1582 | 676 | 546 | 122 |
| History of Hypertension | 339 | 1205 | 1692 | 669 | 564 | 144 |
| **Thiazolidinediones excluding concomitant metformin use** | **314** | **1602** | **631** | **292** | **548** | **144** |
| Male | 164 | 1172 | 270 | 171 | 319 | 84 |
| Female | 150 | 430 | 361 | 121 | 229 | 60 |
| Age <=20 | NA | 11 | 20 | NA | NA | NA |
| Age 21-60 | 73 | 1345 | 442 | -5 | 140 | 41 |
| Age 61-80 | 189 | 246 | 146 | 217 | 320 | 80 |

| | | | | | | |
|---|---|---|---|---|---|---|
| HbA1c > 8% | -5 | NA | NA | NA | 13 | -5 |
| History of MASLD | -5 | NA | 11 | -5 | 5 | NA |
| Renal Impairment not on Dialysis | 52 | 192 | 163 | 111 | 92 | 27 |
| Renal Impairment on Dialysis | -5 | -5 | 10 | -5 | 12 | -5 |
| Obese | 114 | 501 | 294 | 83 | 144 | 52 |
| History of DKA | NA | 7 | 22 | -5 | -5 | -5 |
| History of Diabetic Retinopathy | -5 | NA | 18 | -5 | 12 | -5 |
| History of Hyperlipidemia | 138 | 1133 | 356 | 203 | 212 | 68 |
| History of Hypertension | 163 | 820 | 408 | 209 | 238 | 84 |
| **Meglitinide** | **511** | **778** | **170** | **270** | **471** | **16** |
| Male | 262 | 530 | 78 | 143 | 247 | 9 |
| Female | 249 | 248 | 92 | 127 | 224 | 7 |
| Age <=20 | -5 | 6 | -5 | NA | NA | NA |
| Age 21-60 | 112 | 579 | 113 | -5 | 110 | -5 |
| Age 61-80 | 283 | 193 | 45 | 177 | 277 | 7 |
| HbA1c > 8% | 21 | NA | NA | NA | 41 | NA |
| History of MASLD | -5 | NA | -5 | NA | -5 | NA |
| Renal Impairment not on Dialysis | 137 | 165 | 66 | 126 | 125 | 6 |
| Renal Impairment on Dialysis | 22 | 14 | 8 | 6 | 25 | -5 |
| Obese | 211 | 159 | 90 | 84 | 152 | -5 |
| History of DKA | -5 | 24 | 8 | 10 | 7 | NA |
| History of Diabetic Retinopathy | 14 | NA | 10 | 10 | 14 | NA |
| History of Hyperlipidemia | 257 | 389 | 109 | 219 | 249 | 13 |
| History of Hypertension | 313 | 345 | 129 | 220 | 240 | 12 |
| **Meglitinides excluding concomitant metformin use** | **324** | **500** | **61** | **146** | **253** | **14** |
| Male | 165 | 333 | 27 | 80 | 138 | 8 |
| Female | 159 | 167 | 34 | 66 | 115 | 6 |
| Age <=20 | -5 | -5 | -5 | NA | NA | NA |
| Age 21-60 | 78 | 357 | 30 | -5 | 44 | -5 |
| Age 61-80 | 173 | 139 | 25 | 91 | 152 | 5 |
| HbA1c > 8% | -5 | NA | NA | NA | 24 | NA |
| History of MASLD | -5 | NA | -5 | NA | NA | NA |
| Renal Impairment not on Dialysis | 84 | 108 | 22 | 74 | 75 | 5 |
| Renal Impairment on Dialysis | 18 | 14 | 6 | -5 | 19 | -5 |
| Obese | 123 | 80 | 21 | 46 | 68 | NA |
| History of DKA | -5 | 11 | -5 | -5 | -5 | NA |
| History of Diabetic Retinopathy | -5 | NA | -5 | 5 | 6 | NA |
| History of Hyperlipidemia | 146 | 220 | 31 | 115 | 116 | 11 |
| History of Hypertension | 183 | 218 | 43 | 116 | 119 | 11 |

**Supplementary Table S3.** Cohort sizes among adults with type 2 diabetes mellitus, stratified by index glucose-lowering drug class and demographic subgroups, across data sources. Counts are reported for all index drug class cohorts, including cohorts defined both with and without exclusion of concomitant metformin use. Subgroup counts are shown within each drug class cohort. For privacy protection, cells reported as “–5” indicate suppressed counts representing between 1 and 4 patients.

| Cohort Name | CUIMC *Mean [min-max] (days)* | JMDC *Mean [min-max] (days)* | MDCD *Mean [min-max] (days)* | MDCR *Mean [min-max] (days)* | STARR OMOP *Mean [min-max] (days)* | WashU *Mean [min-max] (days)* |
|---|---|---|---|---|---|---|
| GLP-1 RAs excluding concomitant metformin use | 304 (1-3214) | 373.81 (6-2771) | 248.63 (1-2357) | 328.99 (2-2368) | 364.31 (30-3709) | 576.87 (1-3419) |
| SGLT2is excluding concomitant metformin use | 261 (1-3171) | 566.28 (1-3552) | 258.01 (1-2637) | 322.81 (1-3207) | 315.97 (30-3700) | 524.91 (1-3045) |
| Sulfonylureas excluding concomitant metformin use | 307 (1-3250) | 490.34 (1-3547) | 338.41 (1-3351) | 391.72 (2-3331) | 524.52 (30-3798) | 712.19 (1-3638) |
| DPP4s excluding concomitant metformin use | 302 (1-3079) | 672.99 (1-3555) | 366.74 (1-2769) | 407.39 (2-3076) | 459.64 (30-3711) | 634.91 (1-3512) |
| Alpha-glucosidase inhibitors | 287 (60-474) | 556.86 (2-3546) | 129.2 (59-285) | 208.5 (88-338) | 30 (30-30) | NA |
| Alpha-glucosidase inhibitors excluding concomitant metformin use | 474 (474-474) | 534.12 (2-3379) | 126.25 (59-285) | 204 (119-289) | 30 (30-30) | NA |
| GIP-GLP-1 RA dual agonists | 103 (17-415) | NA | 110.06 (1-557) | 252.66 (2-859) | 223.52 (30-1048) | 243.08 (5-953) |
| GIP-GLP-1 RA dual agonists excluding concomitant metformin use | 99 (30-413) | NA | 114.61 (2-480) | 263.32 (2-859) | 201.19 (30-943) | 243.42 (9-815) |
| Thiazolidinediones | 257 (1-2942) | 621.33 (1-3517) | 384.82 (1-3154) | 367.28 (4-2866) | 490.33 (30-3815) | 572.56 (1-3501) |
| Thiazolidinediones excluding concomitant metformin use | 279 (1-2784) | 606.94 (2-3472) | 362.99 (4-2929) | 357.15 (18-2058) | 462.09 (30-3257) | 521.53 (1-2558) |
| Meglitinide | 271 (1-2888) | 333.96 (3-3285) | 326.39 (21-2096) | 281.69 (9-1933) | 575.98 (30-3790) | 863.31 (32-2696) |
| Meglitinides excluding concomitant metformin use | 262 (1-2489) | 318.89 (3-3285) | 295.13 (30-1504) | 283.08 (9-1933) | 556.17 (30-3228) | 909.79 (64-2696) |

**Supplementary Table S4.** Mean on-treatment follow-up time among adults with type 2 diabetes mellitus who met all study inclusion criteria, stratified by index glucose-lowering drug class and data source. Values are presented as the mean time with the minimum and maximum observed values shown in parentheses (mean [min–max]).

| Outcome | CUIMC, *n* (%) | JMDC, *n* (%) | MDCD, *n* (%) | MDCR, *n* (%) | STARR OMOP, *n* (%) | WashU, *n* (%) |
|---|---|---|---|---|---|---|
| **Biguanides** | | | | | | |
| Acute Pancreatitis | 75 (0.31) | 81 (0.18) | 1047 (0.71) | 189 (0.44) | 226 (0.68) | 22 (0.43) |
| xHepatic Failure | 67 (0.29) | 27 (0.06) | 385 (0.27) | 96 (0.23) | 235 (0.75) | 19 (0.38) |
| Hypoglycemia | 136 (0.59) | 78 (0.19) | 1484 (1.05) | 176 (0.42) | 178 (0.57) | 22 (0.45) |
| Diabetic Ketoacidosis | 134 (0.56) | 69 (0.16) | 1291 (0.89) | 74 (0.18) | 198 (0.59) | 43 (0.88) |
| Abnormal Weight Gain | 173 (0.66) | -5 (NA) | 2704 (1.9) | 195 (0.47) | 278 (0.83) | 0 (0) |
| Acute Renal Failure | 970 (4.11) | 35 (0.08) | 5678 (3.89) | 1605 (3.84) | 1533 (4.62) | 242 (4.9) |
| Vomiting | 943 (3.97) | 1895 (4.39) | 13264 (9.2) | 1654 (3.88) | 1969 (5.83) | 226 (4.39) |
| Diarrhea | 1100 (4.34) | 193 (0.46) | 12020 (8.19) | 2366 (5.5) | 1858 (5.34) | 212 (3.99) |
| Stroke | 274 (1.2) | 115 (0.28) | 1432 (1.02) | 646 (1.56) | 560 (1.8) | 64 (1.3) |
| Hospitalization with Heart Failure Events | 735 (3.05) | 363 (0.83) | 5050 (3.33) | 1865 (4.35) | 1968 (5.47) | 229 (4.31) |
| Acute Myocardial Infarction | 315 (1.35) | 88 (0.21) | 1633 (1.12) | 716 (1.7) | 855 (2.58) | 52 (1.02) |
| **GLP-1 RAs** | | | | | | |
| Acute Pancreatitis | 14 (0.24) | -5 (NA) | 113 (0.55) | 16 (0.36) | 43 (0.55) | 5 (0.39) |
| Hepatic Failure | 12 (0.2) | -5 (NA) | 39 (0.19) | 10 (0.22) | 41 (0.52) | -5 (NA) |
| Hypoglycemia | 52 (0.88) | 5 (0.25) | 150 (0.73) | 19 (0.43) | 59 (0.75) | 5 (0.39) |
| Diabetic Ketoacidosis | 24 (0.41) | 7 (0.37) | 114 (0.55) | -5 (NA) | 36 (0.46) | 8 (0.63) |
| Abnormal Weight Gain | 84 (1.33) | 0 (0) | 316 (1.54) | 39 (0.89) | 96 (1.17) | 0 (0) |
| Acute Renal Failure | 205 (3.38) | -5 (NA) | 554 (2.69) | 131 (2.96) | 358 (4.31) | 48 (3.72) |
| Vomiting | 258 (4.27) | 72 (3.56) | 1845 (9.2) | 184 (4.14) | 494 (5.99) | 62 (4.63) |
| Diarrhea | 287 (4.62) | 5 (0.25) | 1505 (7.4) | 222 (4.9) | 440 (5.2) | 53 (4) |
| Stroke | 49 (0.83) | -5 (NA) | 115 (0.56) | 30 (0.67) | 80 (1.02) | 10 (0.78) |
| Hospitalization with Heart Failure Events | 170 (2.76) | 28 (1.37) | 516 (2.46) | 143 (3.16) | 415 (4.99) | 48 (3.57) |
| Acute Myocardial Infarction | 53 (0.89) | -5 (NA) | 166 (0.78) | 52 (1.17) | 191 (2.34) | 20 (1.57) |
| **GLP-1 RAs excluding concomitant metformin use** | | | | | | |
| Acute Pancreatitis | -5 (NA) | -5 (NA) | 23 (0.47) | 7 (0.27) | 25 (0.72) | -5 (NA) |
| Hepatic Failure | -5 (NA) | -5 (NA) | 15 (0.31) | 7 (0.27) | 27 (0.78) | -5 (NA) |
| Hypoglycemia | 30 (1.11) | -5 (NA) | 34 (0.7) | 7 (0.27) | 32 (0.93) | -5 (NA) |
| Diabetic Ketoacidosis | -5 (NA) | -5 (NA) | 16 (0.33) | 0 (0) | 17 (0.49) | -5 (NA) |
| Abnormal Weight Gain | 31 (1.16) | 0 (0) | 66 (1.39) | 21 (0.84) | 42 (1.17) | 0 (0) |
| Acute Renal Failure | 111 (3.94) | -5 (NA) | 170 (3.52) | 79 (3.13) | 195 (5.38) | 29 (4.11) |

| | | | | | | |
|---|---|---|---|---|---|---|
| Vomiting | 98 (3.55) | 42 (3.47) | 371 (7.98) | 117 (4.59) | 210 (5.85) | 41 (5.58) |
| Diarrhea | 130 (4.47) | -5 (NA) | 287 (6.02) | 135 (5.14) | 183 (4.96) | 25 (3.36) |
| Stroke | 31 (1.15) | -5 (NA) | 38 (0.78) | 16 (0.62) | 47 (1.37) | 7 (0.98) |
| Hospitalization with Heart Failure Events | 84 (2.96) | 19 (1.45) | 163 (3.27) | 89 (3.47) | 226 (6.3) | 31 (4.13) |
| Acute Myocardial Infarction | 26 (0.96) | -5 (NA) | 50 (0.99) | 32 (1.26) | 118 (3.26) | 10 (1.4) |
| **SGLT-2i** | | | | | | |
| Acute Pancreatitis | 17 (0.33) | 93 (0.16) | 97 (0.64) | 15 (0.21) | 35 (0.48) | 6 (0.48) |
| Hepatic Failure | 23 (0.45) | 44 (0.08) | 65 (0.43) | 31 (0.43) | 87 (1.24) | 13 (1.05) |
| Hypoglycemia | 27 (0.53) | 79 (0.14) | 103 (0.68) | 44 (0.61) | 51 (0.72) | 7 (0.56) |
| Diabetic Ketoacidosis | 28 (0.53) | 97 (0.17) | 114 (0.76) | 12 (0.17) | 62 (0.86) | 8 (0.65) |
| Abnormal Weight Gain | 19 (0.37) | -5 (NA) | 144 (0.94) | 37 (0.51) | 34 (0.44) | 0 (0) |
| Acute Renal Failure | 257 (5.22) | 89 (0.16) | 754 (5.3) | 603 (8.99) | 575 (8.34) | 73 (6.04) |
| Vomiting | 195 (3.82) | 2277 (4) | 968 (6.61) | 295 (4.08) | 322 (4.44) | 53 (4.26) |
| Diarrhea | 193 (3.77) | 175 (0.31) | 816 (5.48) | 324 (4.5) | 309 (4.11) | 40 (3.26) |
| Stroke | 57 (1.12) | 178 (0.32) | 172 (1.15) | 143 (2) | 145 (2.07) | 10 (0.81) |
| Hospitalization with Heart Failure Events | 323 (6.91) | 1500 (2.68) | 734 (5.64) | 731 (12.47) | 695 (10.89) | 86 (7.28) |
| Acute Myocardial Infarction | 90 (1.81) | 195 (0.35) | 272 (1.88) | 236 (3.43) | 380 (5.32) | 18 (1.49) |
| **SGLT-2i excluding concomitant metformin use** | | | | | | |
| Acute Pancreatitis | 10 (0.43) | 70 (0.16) | 30 (0.65) | 11 (0.23) | 18 (0.58) | -5 (NA) |
| Hepatic Failure | 16 (0.7) | 35 (0.08) | 40 (0.86) | 26 (0.54) | 58 (1.9) | 11 (1.64) |
| Hypoglycemia | 11 (0.48) | 63 (0.14) | 47 (1.01) | 32 (0.67) | 26 (0.84) | 5 (0.74) |
| Diabetic Ketoacidosis | 12 (0.48) | 63 (0.15) | 23 (0.49) | -5 (NA) | 34 (1.07) | -5 (NA) |
| Abnormal Weight Gain | -5 (NA) | -5 (NA) | 51 (1.1) | 29 (0.61) | 10 (0.32) | 0 (0) |
| Acute Renal Failure | 154 (7.23) | 76 (0.17) | 353 (8.84) | 500 (11.59) | 360 (12.45) | 53 (8.35) |
| Vomiting | 87 (3.8) | 1786 (4.01) | 297 (6.64) | 221 (4.63) | 176 (5.66) | 28 (4.05) |
| Diarrhea | 79 (3.41) | 132 (0.3) | 209 (4.55) | 233 (4.88) | 164 (5.01) | 22 (3.29) |
| Stroke | 35 (1.54) | 151 (0.35) | 77 (1.67) | 107 (2.26) | 91 (3) | 6 (0.89) |
| Hospitalization with Heart Failure Events | 196 (10.05) | 1354 (3.11) | 348 (11.29) | 631 (18) | 443 (18.25) | 63 (10.39) |
| Acute Myocardial Infarction | 55 (2.49) | 156 (0.36) | 143 (3.36) | 200 (4.46) | 222 (7.4) | 14 (2.15) |
| **Sulfonylurea** | | | | | | |
| Acute Pancreatitis | 15 (0.26) | 5 (0.12) | 296 (0.92) | 35 (0.39) | 81 (0.7) | 13 (0.73) |
| Hepatic Failure | 35 (0.61) | 5 (0.12) | 120 (0.39) | 43 (0.49) | 133 (1.19) | 6 (0.34) |

| | | | | | | |
|---|---|---|---|---|---|---|
| Hypoglycemia | 78 (1.35) | 13 (0.31) | 651 (2.13) | 228 (2.59) | 169 (1.5) | 26 (1.45) |
| Diabetic Ketoacidosis | 32 (0.56) | 27 (0.65) | 327 (1.06) | 13 (0.15) | 89 (0.77) | 12 (0.68) |
| Abnormal Weight Gain | 10 (NA) | 0 (0) | 409 (1.29) | 64 (0.72) | 71 (0.6) | 0 (0) |
| Acute Renal Failure | 392 (6.88) | 8 (0.19) | 1673 (5.35) | 672 (7.78) | 887 (7.5) | 147 (7.87) |
| Vomiting | 179 (3.05) | 166 (3.65) | 2590 (8.44) | 481 (5.46) | 703 (5.88) | 76 (4.01) |
| Diarrhea | 216 (3.5) | -5 (NA) | 2356 (7.51) | 519 (5.85) | 637 (5.33) | 82 (4.49) |
| Stroke | 78 (1.36) | 14 (0.34) | 379 (1.24) | 203 (2.33) | 287 (2.57) | 39 (2.19) |
| Hospitalization with Heart Failure Events | 251 (4.3) | 48 (1.08) | 1398 (4.35) | 710 (8.04) | 1054 (8.69) | 146 (7.42) |
| Acute Myocardial Infarction | 109 (1.9) | 10 (0.24) | 451 (1.46) | 237 (2.66) | 390 (3.37) | 31 (1.74) |
| **Sulfonylurea excluding concomitant metformin use** | | | | | | |
| Acute Pancreatitis | -5 (NA) | -5 (NA) | 72 (0.87) | 13 (0.4) | 32 (0.67) | 6 (0.65) |
| Hepatic Failure | 17 (0.66) | -5 (NA) | 48 (0.62) | 18 (0.55) | 81 (1.76) | -5 (NA) |
| Hypoglycemia | 42 (1.63) | 13 (0.45) | 207 (2.66) | 101 (3.1) | 79 (1.7) | 15 (1.62) |
| Diabetic Ketoacidosis | 17 (0.66) | 25 (0.87) | 78 (0.98) | -5 (NA) | 35 (0.73) | 8 (0.86) |
| Abnormal Weight Gain | -5 (NA) | 0 (0) | 97 (1.21) | 25 (0.77) | 29 (0.6) | 0 (0) |
| Acute Renal Failure | 201 (7.84) | 7 (0.24) | 558 (7.23) | 336 (10.71) | 460 (9.65) | 97 (9.77) |
| Vomiting | 82 (3.19) | 111 (3.55) | 595 (7.76) | 176 (5.43) | 272 (5.57) | 42 (4.45) |
| Diarrhea | 89 (3.4) | -5 (NA) | 502 (6.37) | 169 (5.15) | 280 (5.77) | 48 (4.98) |
| Stroke | 40 (1.55) | 9 (0.32) | 118 (1.53) | 81 (2.52) | 131 (2.84) | 27 (2.93) |
| Hospitalization with Heart Failure Events | 141 (5.38) | 30 (0.95) | 470 (6.06) | 325 (10.18) | 556 (11.31) | 103 (9.93) |
| Acute Myocardial Infarction | 55 (2.15) | 6 (0.21) | 136 (1.73) | 100 (3.09) | 185 (3.92) | 23 (2.5) |
| **DPP-4i** | | | | | | |
| Acute Pancreatitis | 17 (0.26) | 126 (0.21) | 106 (0.82) | 28 (0.53) | 27 (0.61) | 0 (0) |
| Hepatic Failure | 31 (0.48) | 87 (0.15) | 45 (0.36) | 25 (0.47) | 47 (1.07) | -5 (NA) |
| Hypoglycemia | 47 (0.73) | 121 (0.21) | 123 (0.99) | 38 (0.72) | 29 (0.65) | 5 (0.83) |
| Diabetic Ketoacidosis | 44 (0.68) | 160 (0.27) | 110 (0.86) | 13 (0.25) | 32 (0.66) | -5 (NA) |
| Abnormal Weight Gain | -5 (NA) | -5 (NA) | 178 (1.4) | 22 (0.41) | 20 (0.43) | 0 (0) |
| Acute Renal Failure | 441 (6.82) | 83 (0.14) | 764 (5.97) | 377 (7.26) | 284 (6.09) | 48 (7.55) |
| Vomiting | 257 (3.8) | 2905 (4.81) | 1216 (9.58) | 250 (4.73) | 258 (5.58) | 30 (5.03) |
| Diarrhea | 283 (4.05) | 199 (0.32) | 1107 (8.55) | 298 (5.55) | 214 (4.39) | 30 (5.05) |
| Stroke | 83 (1.29) | 216 (0.37) | 163 (1.31) | 111 (2.12) | 90 (2.05) | 13 (2.17) |
| Hospitalization with Heart Failure Events | 325 (5.01) | 893 (1.45) | 656 (4.96) | 368 (7.05) | 369 (7.43) | 40 (6.24) |

| | | | | | | |
|---|---|---|---|---|---|---|
| Acute Myocardial Infarction | 117 (1.79) | 195 (0.32) | 177 (1.41) | 130 (2.45) | 147 (3.03) | 8 (1.34) |
| **DPP-4i excluding concomitant metformin use** | | | | | | |
| Acute Pancreatitis | -5 (NA) | 109 (0.22) | 21 (0.96) | 11 (0.63) | 10 (0.61) | 0 (0) |
| Hepatic Failure | 11 (0.5) | 77 (0.16) | 12 (0.55) | 8 (0.46) | 22 (1.35) | -5 (NA) |
| Hypoglycemia | 19 (0.85) | 100 (0.21) | 30 (1.37) | 14 (0.81) | 18 (1.1) | 5 (1.62) |
| Diabetic Ketoacidosis | 17 (0.77) | 139 (0.29) | 14 (0.6) | -5 (NA) | 12 (0.67) | -5 (NA) |
| Abnormal Weight Gain | -5 (NA) | -5 (NA) | 39 (1.74) | 10 (0.57) | 7 (0.43) | 0 (0) |
| Acute Renal Failure | 201 (9.26) | 71 (0.15) | 219 (10.19) | 174 (10.46) | 146 (8.52) | 38 (11.95) |
| Vomiting | 97 (4.07) | 2415 (4.94) | 192 (8.59) | 94 (5.43) | 101 (5.97) | 17 (5.61) |
| Diarrhea | 105 (4.39) | 158 (0.31) | 177 (7.79) | 96 (5.55) | 79 (4.36) | 14 (4.61) |
| Stroke | 33 (1.5) | 180 (0.38) | 35 (1.61) | 37 (2.18) | 50 (3.08) | 8 (2.62) |
| Hospitalization with Heart Failure Events | 161 (7.46) | 765 (1.54) | 193 (8.77) | 146 (9.11) | 190 (10.6) | 35 (10.69) |
| Acute Myocardial Infarction | 52 (2.33) | 159 (0.32) | 61 (2.73) | 48 (2.84) | 76 (4.33) | 5 (1.63) |
| **Alpha-glucosidase inhibitors** | | | | | | |
| Acute Pancreatitis | 0 (0) | -5 (NA) | 0 (0) | 0 (0) | 0 (0) | 0 (NA) |
| Hepatic Failure | 0 (0) | -5 (NA) | 0 (0) | 0 (0) | 0 (0) | 0 (NA) |
| Hypoglycemia | 0 (0) | 8 (0.46) | -5 (NA) | 0 (0) | 0 (0) | 0 (NA) |
| Diabetic Ketoacidosis | 0 (0) | -5 (NA) | 0 (0) | 0 (0) | 0 (0) | 0 (NA) |
| Abnormal Weight Gain | 0 (0) | -5 (NA) | 0 (0) | -5 (NA) | 0 (0) | 0 (NA) |
| Acute Renal Failure | 0 (0) | 0 (0) | 0 (0) | 0 (0) | 0 (0) | 0 (NA) |
| Vomiting | 0 (0) | 72 (4.01) | 0 (0) | 0 (0) | 0 (0) | 0 (NA) |
| Diarrhea | -5 (NA) | -5 (NA) | 0 (0) | 0 (0) | 0 (0) | 0 (NA) |
| Stroke | 0 (0) | 5 (0.29) | 0 (0) | 0 (0) | 0 (0) | 0 (NA) |
| Hospitalization with Heart Failure Events | 0 (0) | 22 (1.27) | 0 (0) | -5 (NA) | 0 (0) | 0 (NA) |
| Acute Myocardial Infarction | 0 (0) | 5 (0.29) | 0 (0) | 0 (0) | 0 (0) | 0 (NA) |
| **Alpha-glucosidase inhibitors excluding concomitant metformin use** | | | | | | |
| Acute Pancreatitis | 0 (0) | -5 (NA) | 0 (0) | 0 (0) | 0 (0) | 0 (NA) |
| Hepatic Failure | 0 (0) | -5 (NA) | 0 (0) | 0 (0) | 0 (0) | 0 (NA) |
| Hypoglycemia | 0 (0) | -5 (NA) | -5 (NA) | 0 (0) | 0 (0) | 0 (NA) |
| Diabetic Ketoacidosis | 0 (0) | -5 (NA) | 0 (0) | 0 (0) | 0 (0) | 0 (NA) |
| Abnormal Weight Gain | 0 (0) | -5 (NA) | 0 (0) | 0 (0) | 0 (0) | 0 (NA) |
| Acute Renal Failure | 0 (0) | 0 (0) | 0 (0) | 0 (0) | 0 (0) | 0 (NA) |

| | | | | | | |
|---|---|---|---|---|---|---|
| Vomiting | 0 (0) | 48 (3.56) | 0 (0) | 0 (0) | 0 (0) | 0 (NA) |
| Diarrhea | -5 (NA) | -5 (NA) | 0 (0) | 0 (0) | 0 (0) | 0 (NA) |
| Stroke | 0 (0) | -5 (NA) | 0 (0) | 0 (0) | 0 (0) | 0 (NA) |
| Hospitalization with Heart Failure Events | 0 (0) | 19 (1.43) | 0 (0) | -5 (NA) | 0 (0) | 0 (NA) |
| Acute Myocardial Infarction | 0 (0) | -5 (NA) | 0 (0) | 0 (0) | 0 (0) | 0 (NA) |
| **GIP-GLP-1 RA dual agonists** | | | | | | |
| Acute Pancreatitis | 0 (0) | 0 (NA) | 0 (0) | -5 (NA) | 0 (0) | -5 (NA) |
| Hepatic Failure | 0 (0) | 0 (NA) | 0 (0) | -5 (NA) | -5 (NA) | -5 (NA) |
| Hypoglycemia | 0 (0) | 0 (NA) | -5 (NA) | 7 (0.73) | -5 (NA) | -5 (NA) |
| Diabetic Ketoacidosis | 0 (0) | 0 (NA) | -5 (NA) | 0 (0) | -5 (NA) | 0 (0) |
| Abnormal Weight Gain | -5 (NA) | 0 (NA) | -5 (NA) | 6 (0.64) | -5 (NA) | 0 (0) |
| Acute Renal Failure | -5 (NA) | 0 (NA) | -5 (NA) | 21 (2.21) | 13 (1.72) | 6 (2.01) |
| Vomiting | -5 (NA) | 0 (NA) | 14 (2.68) | 20 (2.11) | 21 (2.85) | -5 (NA) |
| Diarrhea | -5 (NA) | 0 (NA) | 11 (2.09) | 36 (3.8) | 15 (2.03) | 10 (3.31) |
| Stroke | 0 (0) | 0 (NA) | -5 (NA) | -5 (NA) | 5 (0.66) | 0 (0) |
| Hospitalization with Heart Failure Events | 0 (0) | 0 (NA) | -5 (NA) | 24 (2.54) | 21 (2.69) | 5 (1.68) |
| Acute Myocardial Infarction | -5 (NA) | 0 (NA) | 0 (0) | -5 (NA) | 11 (1.47) | -5 (NA) |
| **GIP-GLP-1 RA dual agonists excluding concomitant metformin use** | | | | | | |
| Acute Pancreatitis | 0 (0) | 0 (NA) | 0 (0) | -5 (NA) | 0 (0) | 0 (0) |
| Hepatic Failure | 0 (0) | 0 (NA) | 0 (0) | -5 (NA) | -5 (NA) | -5 (NA) |
| Hypoglycemia | 0 (0) | 0 (NA) | -5 (NA) | 5 (0.74) | -5 (NA) | -5 (NA) |
| Diabetic Ketoacidosis | 0 (0) | 0 (NA) | -5 (NA) | 0 (0) | 0 (0) | 0 (0) |
| Abnormal Weight Gain | -5 (NA) | 0 (NA) | -5 (NA) | 5 (0.76) | -5 (NA) | 0 (0) |
| Acute Renal Failure | -5 (NA) | 0 (NA) | -5 (NA) | 15 (2.24) | 5 (1.47) | -5 (NA) |
| Vomiting | -5 (NA) | 0 (NA) | 7 (2.69) | 17 (2.55) | 9 (2.75) | -5 (NA) |
| Diarrhea | -5 (NA) | 0 (NA) | 7 (2.65) | 31 (4.65) | 5 (1.51) | -5 (NA) |
| Stroke | 0 (0) | 0 (NA) | -5 (NA) | -5 (NA) | -5 (NA) | 0 (0) |
| Hospitalization with Heart Failure Events | 0 (0) | 0 (NA) | -5 (NA) | 17 (2.56) | 13 (3.61) | -5 (NA) |
| Acute Myocardial Infarction | -5 (NA) | 0 (NA) | 0 (0) | -5 (NA) | 5 (1.48) | -5 (NA) |
| **Thiazolidinediones** | | | | | | |
| Acute Pancreatitis | -5 (NA) | -5 (NA) | 20 (0.8) | -5 (NA) | 5 (NA) | -5 (NA) |
| Hepatic Failure | -5 (NA) | -5 (NA) | 11 (0.44) | -5 (NA) | 11 (0.97) | -5 (NA) |

| | | | | | | |
|---|---|---|---|---|---|---|
| Hypoglycemia | -5 (NA) | 0 (0) | 17 (0.68) | 8 (0.9) | 8 (0.7) | -5 (NA) |
| Diabetic Ketoacidosis | -5 (NA) | -5 (NA) | 23 (0.89) | -5 (NA) | 6 (0.53) | -5 (NA) |
| Abnormal Weight Gain | 0 (0) | 0 (0) | 54 (2.14) | 7 (0.78) | 8 (0.7) | 0 (0) |
| Acute Renal Failure | 29 (5.05) | -5 (NA) | 108 (4.25) | 37 (4.16) | 50 (4.37) | 18 (7.59) |
| Vomiting | 15 (2.62) | 95 (4.18) | 220 (8.68) | 39 (4.34) | 44 (3.84) | 11 (4.64) |
| Diarrhea | 22 (3.66) | 10 (0.44) | 189 (7.45) | 45 (5.06) | 38 (3.02) | 10 (3.78) |
| Stroke | -5 (NA) | -5 (NA) | 31 (1.24) | 11 (1.24) | 21 (1.87) | 0 (0) |
| Hospitalization with Heart Failure Events | 20 (3.51) | 13 (0.52) | 102 (3.89) | 36 (4.15) | 84 (7.2) | 16 (6.36) |
| Acute Myocardial Infarction | -5 (NA) | -5 (NA) | 31 (1.24) | 13 (1.47) | 23 (2.04) | -5 (NA) |
| **Thiazolidinediones excluding concomitant metformin use** | | | | | | |
| Acute Pancreatitis | 0 (0) | -5 (NA) | 7 (1.13) | -5 (NA) | 0 (0) | 0 (0) |
| Hepatic Failure | -5 (NA) | -5 (NA) | -5 (NA) | -5 (NA) | 7 (1.28) | -5 (NA) |
| Hypoglycemia | -5 (NA) | 0 (0) | 6 (0.97) | -5 (NA) | 6 (1.09) | -5 (NA) |
| Diabetic Ketoacidosis | 0 (0) | -5 (NA) | 10 (1.61) | -5 (NA) | -5 (NA) | 0 (0) |
| Abnormal Weight Gain | 0 (0) | 0 (0) | 15 (2.26) | -5 (NA) | 5 (0.91) | 0 (0) |
| Acute Renal Failure | 17 (5.48) | -5 (NA) | 40 (6.3) | 14 (4.63) | 31 (5.74) | 13 (9.22) |
| Vomiting | -5 (NA) | 75 (4.73) | 52 (8.18) | 10 (3.51) | 26 (4.61) | 6 (4.26) |
| Diarrhea | 12 (3.9) | 7 (0.44) | 51 (8.17) | 12 (4.17) | 22 (3.68) | -5 (NA) |
| Stroke | -5 (NA) | -5 (NA) | 8 (1.28) | -5 (NA) | 11 (2.03) | 0 (0) |
| Hospitalization with Heart Failure Events | 13 (4.25) | 9 (0.56) | 42 (6.21) | 8 (2.86) | 44 (7.68) | 14 (9.29) |
| Acute Myocardial Infarction | -5 (NA) | -5 (NA) | 11 (1.77) | -5 (NA) | 14 (2.57) | -5 (NA) |
| **Meglitinide** | | | | | | |
| Acute Pancreatitis | 0 (0) | -5 (NA) | -5 (NA) | -5 (NA) | -5 (NA) | 0 (0) |
| Hepatic Failure | -5 (NA) | 0 (0) | 0 (0) | 0 (0) | 7 (1.49) | 0 (0) |
| Hypoglycemia | 11 (2.16) | -5 (NA) | 7 (4.14) | 5 (1.88) | 5 (1.06) | 0 (0) |
| Diabetic Ketoacidosis | -5 (NA) | 0 (0) | -5 (NA) | 0 (0) | -5 (NA) | 0 (0) |
| Abnormal Weight Gain | 0 (0) | 0 (0) | -5 (NA) | -5 (NA) | -5 (NA) | 0 (0) |
| Acute Renal Failure | 35 (7.16) | -5 (NA) | 8 (4.7) | 21 (8.82) | 34 (7.19) | -5 (NA) |
| Vomiting | 17 (3.4) | 30 (4.1) | 12 (7.55) | 9 (3.47) | 42 (8.3) | 0 (0) |
| Diarrhea | 35 (6.59) | 0 (0) | 18 (11.32) | 14 (5.56) | 31 (6.47) | 0 (0) |
| Stroke | -5 (NA) | -5 (NA) | -5 (NA) | -5 (NA) | 14 (2.98) | 0 (0) |
| Hospitalization with Heart Failure Events | 20 (4.14) | 9 (1.2) | 12 (6.62) | 20 (8.33) | 44 (8.67) | -5 (NA) |

| | | | | | | |
|---|---|---|---|---|---|---|
| Acute Myocardial Infarction | 15 (2.97) | 0 (0) | -5 (NA) | -5 (NA) | 14 (2.77) | 0 (0) |
| **Meglitinide excluding concomitant metformin use** | | | | | | |
| Acute Pancreatitis | 0 (0) | -5 (NA) | -5 (NA) | -5 (NA) | -5 (NA) | 0 (0) |
| Hepatic Failure | -5 (NA) | 0 (0) | 0 (0) | 0 (0) | -5 (NA) | 0 (0) |
| Hypoglycemia | -5 (NA) | -5 (NA) | -5 (NA) | -5 (NA) | -5 (NA) | 0 (0) |
| Diabetic Ketoacidosis | -5 (NA) | 0 (0) | -5 (NA) | 0 (0) | -5 (NA) | 0 (0) |
| Abnormal Weight Gain | 0 (0) | 0 (0) | 0 (0) | -5 (NA) | -5 (NA) | 0 (0) |
| Acute Renal Failure | 24 (7.79) | -5 (NA) | 5 (NA) | 13 (10.48) | 22 (8.98) | -5 (NA) |
| Vomiting | 12 (3.82) | 17 (3.88) | -5 (NA) | -5 (NA) | 29 (10.25) | 0 (0) |
| Diarrhea | 23 (6.62) | 0 (0) | -5 (NA) | 6 (4.38) | 16 (5.98) | 0 (0) |
| Stroke | -5 (NA) | -5 (NA) | -5 (NA) | -5 (NA) | 9 (3.57) | 0 (0) |
| Hospitalization with Heart Failure Events | 14 (4.58) | 7 (1.46) | 6 (9.26) | 14 (11.02) | 26 (9.96) | -5 (NA) |
| Acute Myocardial Infarction | 12 (3.75) | 0 (0) | -5 (NA) | -5 (NA) | 10 (3.56) | 0 (0) |

**Supplementary Table S5.** Outcome counts and incidence proportions (per 100 persons) among adults with type 2 diabetes mellitus who met all study inclusion criteria, stratified by index glucose-lowering drug class and data source. Values are reported as the number of patients experiencing each outcome and the corresponding incidence proportion per 100 persons during observed follow-up. Drug exposure was defined based on the first observed initiation of a medication from one of the prespecified glucose-lowering drug classes. To preserve patient privacy, outcome counts between 1 and 4 were suppressed and displayed as “–5”; incidence proportions were therefore not calculated or reported for these cells.
